# 3D chromatin-based variant-to-gene maps across 57 human cell types reveal the cellular and genetic architecture of autoimmune disease susceptibility

**DOI:** 10.1101/2024.08.12.24311676

**Authors:** Khanh B. Trang, Prabhat Sharma, Laura Cook, Zachary Mount, Rajan M. Thomas, Nikhil N. Kulkarni, Emylette Cruz Cabrera, Suzanna Rachimi, Matthew C. Pahl, James A. Pippin, Chun Su, Klaus H. Kaestner, Joan M. O’Brien, Yadav Wagley, Kurt D. Hankenson, Ashley Jermusyk, Jason W. Hoskins, Laufey T. Amundadottir, Mai Xu, Kevin M. Brown, Stewart A. Anderson, Wenli Yang, Paul M. Titchenell, Patrick Seale, Babette S. Zemel, Alessandra Chesi, Neil Romberg, Megan K. Levings, Struan F.A. Grant, Andrew D. Wells

## Abstract

Genome-wide association studies (GWAS) have identified genetic links to autoimmune disorders, but lack detail on causal elements. We generated 3D genomic datasets of promoter-focused Capture-C, Hi-C, ATAC-seq, and RNA-seq across 57 human cell types integrated with GWAS of 16 autoimmune traits. These data allowed us to map disease-associated variants to their effector genes and identify impacted cell types more effectively than using 1D genomic features or eQTL approaches. Most variants implicated by 3D cis-regulatory architectures are trait-specific, while half the target genes are shared across multiple disorders and cell types, leading to enrichment of similar biological networks. This indicates complex genetic diversity converges on shared targets, yet unique pathways were identified offering avenues for targeted therapies. We pharmacologically validated squalene synthase, a cholesterol biosynthetic enzyme encoded by the *FDFT1* gene implicated by our approach and eQTL in multiple sclerosis and systemic lupus erythematosus, as a novel immunomodulatory drug target controlling T cell inflammatory cytokine production and aiding B cell antibody production in a human lymphoid organoid model. These data offer a comprehensive resource for understanding gene cis-regulatory mechanisms, and the analyses shed light on how autoimmune-associated variants regulate gene expression, function, and pathology across diverse tissues and cell types.

## INTRODUCTION

Autoimmune diseases are a group of complex disorders arising from dysregulated inflammatory responses that damage self-tissues and exhibit both shared and unique etiologies. Rare, naturally occurring or engineered mutations in patients or animal models have identified a handful of genes and cell types that contribute to the development and progression of disease, but in the majority of cases, common genetic variation between humans drives autoimmune disease susceptibility. The polygenic and pleiotropic nature of autoimmunity poses serious challenges to comprehensive identification of causal variants, effector genes, effector cell types, novel therapies, and clinical management strategies.

Examples of genes implicated by both common and rare genetic variation to date are *CTLA4,* with known roles in rheumatoid arthritis (RA), type 1 diabetes (T1D), and Hashimoto’s thyroiditis (HT)(1–3), *PTPN22* in systemic lupus erythematosus (SLE), T1D, RA and Grave’s disease (GD)(4,5), and *IL23R* in ulcerative colitis (UC), Crohn’s disease (CRO), SLE, RA and psoriasis (PSO)(6–8). These factors control the activation, proliferation, and function of pathogenic T helper cells and regulatory T cells, which are cell types known to mediate or control inflammation in autoimmune animal models and in humans. Identifying additional effector genes for these traits will be important for understanding disease pathology and revealing new therapeutic approaches.

The majority of autoimmune disease-associated variants have been uncovered with GWAS. Most GWAS signals are non-coding in nature, commonly located far from their effector genes, and their contribution to disease remains unclear. The human genome is packed and folded tightly into chromatin such that only ∼1% of DNA participates in gene regulation(9), and these non-coding *cis*-regulatory elements (cRE) can influence gene expression by ‘looping’ to physically contact distant gene promoters(10).

In this study, we leveraged existing autoimmune GWAS summary statistics and our own matched ATAC-seq, RNA-seq, and high-resolution chromosome conformation capture data from 57 diverse human cell types to implicate causal regulatory autoimmune single nucleotide polymorphisms (SNPs) in open chromatin and to comprehensively map these variants to the genes they likely control. This analysis revealed shared, trait-specific, and cell type-specific gene regulatory networks likely influenced by autoimmune disease-associated genetic variation, including ‘druggable’ targets with no prior known role in immune regulation or autoimmune disease. An example we took forward to functional validation efforts is the *FDFT1* gene encoding a lipid biosynthetic enzyme, which exhibits significantly increased expression in multiple immune cell types from RA and SLE patients compared to healthy subjects, and we show regulates cytokine production and B cell helper function by human T cells.

## MATERIAL AND METHODS

### Data and resource

**Table S1** lists all datasets, including new datasets and datasets used in this study that were previously published. The original published studies provided configurations and technical details in ATAC-seq, Hi-C, and Capture-C ***library generation***.

### Immune cell isolation

The pDC, cDC1, and NK cells were purified from the apheresis products obtained from healthy, screened, consented human donors through University of Pennsylvania Human Immunology Core (HIC). Surface markers used for sorting were: pDC = CD3^neg^ CD14^neg^ CD16^neg^ CD19^neg^ CD20^neg^ CD56^neg^ HLADR+ CD11c^neg^, cDC2 = CD3^neg^ CD14^neg^ CD16^neg^ CD19^neg^ CD20^neg^ CD56^neg^ HLADR+ CD11c+ CD1c+, NK = CD45+ CD3^neg^ CD56+. Gating strategies are included in **Supplemental File 1**. For T cell subsets, peripheral blood in the form of buffy coat products was obtained from anonymized healthy adults of sex and age ranges indicated in **Table S2** via Canadian Blood Services (Vancouver BC, Canada) with informed consent and ethical approval from The University of British Columbia Clinical and Canadian Blood Service Research Ethics Boards (H18-02553). CD4^+^ T cells were obtained by incubation with CD4^+^ RosetteSep enrichment cocktail (Stemcell Technologies) before centrifugation over Ficoll-Paque (GE Healthcare). *Ex vivo* CD4^+^ T cell subsets were isolated to >95% purity by FACS with the following surface markers: Treg = CD25^+^ CD127^low^; Th1 = CD25^neg^ CXCR3^+^ CCR6^neg^ CCR4^neg^; Th2 = CD25^neg^ CXCR3^neg^ CCR6^neg^ CCR4^+^ and Th17 = CD25^neg^ CXCR3^neg^ CCR6^+^ CCR4^+^. Tr1 cells (CD4^+^ CD45RA^neg^ IL10^+^) were isolated following 16 hours anti-CD3/CD28 stimulation using IL-10 secretion capture assay as previously described(11). The mAbs used are listed in **Table S2**. T cell subsets were expanded as previously described(12). Briefly, artificial APCs expressing CD32, CD80, and CD58 were irradiated (7500 rad), loaded with anti-CD3 mAb (OKT3, 100 ng/mL), and used to stimulate a 1:1 ratio of sorted T cells in the presence of recombinant human IL-2 (100U/mL for non-Tregs and 1000U/mL for Tregs) for two weeks, with media and IL-2 replenished every 3-4 days. At day 14, after cells had returned to a resting state, aliquots of 1-2 × 10^5^ cells were stored at −80°C as dry cell pellets (for DNA extraction) or in 100μL RNALater (Sigma-Aldrich, for RNA extraction). Then 10 × 10^6^ cells were set aside for lysis/fixation and remaining cells were cryopreserved. Gating strategies and cell characterization data are included in **Supplemental File 1**.

### Purification and stimulation of CD4+ T cells from human tonsil

Fresh tonsils were obtained as discarded surgical waste from de-identified immune-competent children undergoing tonsillectomy to address airway obstruction or a history of recurrent tonsillitis. These studies were approved by The Children’s Hospital of Philadelphia Institutional Review Board as non-human subject research. The mean age of donors was 5.6 years (range 3-16 years) and 75% were male. Tonsillar mononuclear cells were isolated from tissues by mechanical disruption (tonsils were minced and pressed through a 70-micron cell screen) followed by Ficoll-Paque centrifugation. Total CD4+ T cells were purified using EasySep^TM^ human CD4+ T cell isolation kit I (STEM cells Technologies, cat#17555) by immunomagnetic negative selection as per manufacturer’s protocol. Isolated untouched, highly purified (93-98%) human CD4 T cells were activated using anti-CD3+anti-CD28 Dynabeads (1:1, Thermofisher scientific, cat # 11161D) for 8-24 hours. IL-2, IL-17A, IL-10, IL-4, TNF, and IFNg in supernatants were measured using LEGENDplex^TM^ human cytokine capture beads (Biolegend) on a Cytoflex flow cytometer (Beckman).

### Tonsillar organoid preparation and staining

Excised tonsils from five de-identified immunocompetent patients were diced and strained through a 70-μm filter. A single cell suspension of tonsillar mononuclear cells (MNCs) was created with Ficoll density gradient separation. Once isolated, MNC were counted and resuspended at a concentration of 6 × 10^7^ cells per ml in organoid media (RPMI with L-glutamine, 10% FBS, 2 mM L-glutamine, 1X penicillin-streptomycin, 1 mM sodium pyruvate, 1X MEM non-essential amino acids and 10 mM HEPES buffer). MNC suspensions were transferred to permeable transwells (0.4-μm pore, 12-mm diameter; Millipore, 100µl per well). Transwells were inserted into standard 12-well polystyrene plates containing 1 ml of additional organoid media, supplemented with 1 μg/ml of recombinant human B cell activating factor (rBAFF; BioLegend), and treated, or not, with Lapaquistat in dimethyl sulfoxide (DMSO). Plain DMSO was added to the organoids not treated with Lapaquistat at a concentration of 0.27% (same concentration of DMSO present in the organoids treated with Lapaquistat). Organoids were immunized with the Tenivac (diphtheria-tetanus) vaccine, incubated at 37°C and 5% CO_2_, and organoid media supplemented with rBAFF, with or without Lapaquistat, was replaced at days 2 and 5 of culture. On culture day 7, organoid MNCs were resuspended and stained at 4°C with the anti-human CD38, CD27, CD19, CD45RA, CD45R), CD4, PD1, and CXCR5 (all mAb from BioLegend) and fixable viability dye efluor 780 (Invitrogen). Cells were analyzed on an Aurora D cytometer (Cytek Biosciences) with FlowJo software (TreeStar). Total IgM, IgG, and IgA secretion was measured by ELISA (R&D Systems).

### Cell fixation

Briefly, ∼10^7^ cells were suspended in 10 mL of RPMI medium supplemented with 10% FBS. To fix the cells, 270 μL of 37% formaldehyde was added, and the suspension was incubated for 10 minutes at room temperature on a platform rocker. The fixation reaction was then quenched by adding 1.5 mL of cold 1M glycine (4°C). The fixed cells were centrifuged at 210 × g for 5 minutes at 4°C, and the supernatant was removed. The cell pellets were washed with 10 mL of cold PBS (4°C) and centrifuged again under the same conditions. After washing, the cell pellets were resuspended in 5 mL of cold lysis buffer (10 mM Tris pH 8, 10 mM NaCl, 0.2% NP-40/Igepal, supplemented with a protease inhibitor cocktail) and incubated on ice for 20 minutes. Following incubation, the cells were centrifuged at 680 × g, and the lysis buffer was removed. The cell pellets were then resuspended in 1 mL of fresh lysis buffer, transferred to 1.5 mL Eppendorf tubes, and snap frozen in ethanol/dry ice or liquid nitrogen. The frozen cell pellets were stored at −80°C for subsequent 3C library generation.

### 3C library generation

For each library, 10^7^ fixed cells were thawed at 37°C, followed by centrifugation at room temperature for 5 minutes at 1845 × g. The resulting cell pellet was resuspended in 1 mL of deionized water (dH2O) supplemented with 5 μL of a 200X protease inhibitor cocktail, incubated on ice for 10 minutes, and then centrifuged again. The cell pellet was then resuspended in dH2O to a total volume of 650 μL. A 50 μL aliquot of the cell suspension was set aside for pre-digestion quality control (QC), while the remaining sample was divided into three tubes. Both the pre-digestion control and the samples underwent a pre-digestion incubation in a Thermomixer (BenchMark) with the addition of 0.3% SDS, 1× NEB DpnII restriction buffer, and dH2O, shaking at 1,000 rpm for 1 hour at 37°C. Following this, a 1.7% solution of Triton X-100 was added to each tube, and shaking was continued for another hour. After the pre-digestion incubation, 10 μL of DpnII enzyme (NEB, 50 U/μL) was added to each sample tube (but not the control), and the incubation with shaking continued until the end of the day. An additional 10 μL of DpnII was added to each digestion reaction for an overnight digestion. The next day, a further 10 μL of DpnII was added, and the samples were incubated with shaking for another 2-3 hours. Next, 100 μL from each digestion reaction was pooled into a single 1.5 mL tube and set aside for digestion efficiency QC. The remaining samples were heat inactivated by incubating them at 65°C for 20 minutes in a MultiTherm shaking at 1,000 rpm, followed by cooling on ice for an additional 20 minutes. The digested samples were then ligated by adding 8 μL of T4 DNA ligase (HC ThermoFisher, 30 U/μL) and 1X ligase buffer, followed by overnight incubation at 16°C with shaking at 1,000 rpm in a MultiTherm. The next day, 2 μL more of T4 DNA ligase was added to each sample, and the incubation was continued for a few more hours. The ligated samples were then de-crosslinked overnight at 65°C with Proteinase K (20 mg/mL, Denville Scientific) along with pre-digestion and digestion controls. The following morning, both controls and ligated samples were incubated for 30 minutes at 37°C with RNase A (Millipore), followed by phenol/chloroform extraction and ethanol precipitation at −20°C. The 3C libraries were centrifuged at 1,000 × g for 45 minutes at 4°C to pellet the samples, while the controls were centrifuged at 1845 × g. The pellets were then resuspended in 70% ethanol and centrifuged again as described above. Finally, the pellets of the 3C libraries and controls were resuspended in 300 μL and 20 μL of dH2O, respectively, and stored at −20°C. Sample concentrations were measured using a Qubit fluorometer, and digestion and ligation efficiencies were assessed by gel electrophoresis on a 0.9% agarose gel and quantitative PCR (SYBR Green, Thermo Fisher).

### Promoter Capture-C (PCC) pre-processing and interaction calling

In brief, paired-end reads were pre-processed using HICUP pipeline(13) with bowtie2 as aligner and GRCh37/hg19 for reference genome. Significant promoters interactions were called using unique read pairs from all baits promoter in the reference by CHICAGO(14) pipeline. In addition to analysis of individual fragments (1frag), we also binned four fragments to improve long-distance sensitivity in interactions calling(15). Interactions with CHICAGO score > 5 in either 1-fragment or 4-fragment resolution were considered significant. These interactions were output as *ibed* format (similar to BEDPE format) in which each line represents one physical contact between fragments. Interactions from both resolutions were merged and their genomic coordinates were lifted from GRCh37/hg19 to GRCh38/hg38.

### RNA-seq library generation and sequencing

RNA was isolated from approximately 1 million cells at each stage using Trizol Reagent (Invitrogen), followed by purification with the Direct-zol RNA Miniprep Kit (Zymo Research). To remove any contaminating genomic DNA, the RNA was treated with DNase I. The quality of the purified RNA was assessed using a Bioanalyzer 2100 with the Nano RNA Chip, and only samples with RIN>7 were selected for RNA-seq library preparation. To prepare the RNA samples for sequencing, rRNA was depleted using the QIAseq FastSelect RNA removal kit (Qiagen). The remaining RNA was then processed into RNA-seq libraries using the SMARTer Stranded Total RNA Sample Prep Kit (Takara Bio USA), following the manufacturer’s instructions. Specifically, the purified first-strand cDNA was amplified into RNA-seq libraries using SeqAmp DNA Polymerase along with Forward and Reverse PCR Primers from the Illumina Indexing Primer Set HT. The quality and quantity of the RNA-seq libraries were assessed using the Agilent 2100 Bioanalyzer system and a Qubit fluorometer (Life Technologies). Sequencing was conducted on the NovaSeq 6000 platform at the CHOP Center for Spatial and Functional Genomics.

### RNA-seq preprocessing and expression profiling

The detail configurations, steps, and technical details for each data set are provided in the original studies. In brief, read fragments from fastq files were mapped to genome assembly GRCh37/hg19 or GRCh38/hg38 using **STAR**(16), independently for each replicate and condition. We used GENCODE annotation files for feature annotation and htseq-count for raw read count calculation at each feature. Read counts were transformed into TPM (transcript per million) and normalized internally between replicates/conditions in each individual study. For comparative measurements, we transformed all the expression values into 0-100 scale.

### ATAC-seq library generation and sequencing

A total of 50,000 to 100,000 sorted cells were centrifuged at 550 g for 5 min at 4°C. The cell pellet was washed with cold PBS and resuspended in 50 μL cold lysis buffer (10 mM Tris-HCl, pH 7.4, 10 mM NaCl, 3 mM MgCl2, 0.1% NP-40/IGEPAL CA-630) and immediately centrifuged at 550 g for 10 min at 4°C. Nuclei were resuspended in the Nextera transposition reaction mix (25 μL 2× TD Buffer, 2.5 μL Nextera Tn5 transposase (Illumina Cat #FC-121-1030), and 22.5 μL nuclease free H2O) on ice, then incubated for 45 min at 37°C. The tagmented DNA was then purified using the Qiagen MinElute kit eluted with 10.5 μL Elution Buffer (EB). Ten microliters of purified tagmented DNA was PCR amplified using Nextera primers for 12 cycles to generate each library. PCR reaction was subsequently cleaned up using 1.5× AMPureXP beads (Agencourt), and concentrations were measured by Qubit. Libraries were paired-end sequenced on the Illumina HiSeq 4000 platform (100 bp read length).

### ATAC-seq preprocessing and peaks calling

Open chromatin regions (OCRs) were called using the ENCODE ATAC-seq pipeline (https://github.com/ENCODE-DCC/atac-seq-pipeline). In brief, reads were aligned to GRCh37/hg19 assembly or GRCh38/hg38 assembly genome using bowtie2, duplicates were removed, alignments from all replicates were pooled, narrow peaks were call using ***MACS2***. We lifted all coordinates from GRCh37/hg19 to GRCh38/hg38 to ensure consistency between datasets, using *LiftOver (*https://genome.sph.umich.edu/wiki/LiftOver).

### ATAC-seq transcription factor footprint analysis with RGT toolbox

Bam files of mapped reads from replicates and samples were merged for each cell type. The merged bam files were then used for footprint analysis by ***RGT-HINT***(17) with parameters: *--atac-seq, –paired-end, –organism=hg38*. If bam files were generated on GRCh37/hg19, we performed lift-over to GRCh38/hg38 using ***CrossMap (v0.1.1)***(18) and hg19ToHg38.over.chain.gz file. We then used RGT-MOTIFANALYSIS *matching* to scan each footprint for possible transcription binding sites from HOCOMOCO and JASPAR databases for human only with parameter -- filter “species:sapiens;database:hocomoco,jaspar_vertebrates”. Parameter *–rand-proportion 10* was used to generate random putative binding sites with sizes ten times larger than the input footprints. After performing motif matching, we evaluated which transcription factors were more likely to occur in those footprints than in background regions (generated by the previous command) using RGT-MOTIFANALYSIS *enrichment* with the same filtered databases and default parameters. Output included all the Motif-Predicted Binding Sites (MPBS) that occurred within the identified footprints in each cell type. We overlapped these sites with the loci of all the variants recovered during variant-to-gene analysis.

### Hi-C library preparation

Hi-C library preparation on FACS-sorted cells was conducted using the Arima-HiC kit (Arima Genomics Inc), following the manufacturer’s instructions. In brief, cells were first crosslinked with formaldehyde. After crosslinking, the cells were processed according to the Arima-HiC protocol, which involves digesting chromatin with multiple restriction enzymes. The sequencing libraries were then prepared by shearing the purified, proximally-ligated DNA and selecting DNA fragments ranging from 200 to 600 bp using AmpureXP beads (Beckman Coulter). These size-selected fragments were enriched using Enrichment Beads provided in the Arima-HiC kit and subsequently converted into Illumina-compatible sequencing libraries using the Swift Accel-NGS 2SPlus DNA Library Kit (Swift, 21024) and Swift 2S Indexing Kit (Swift, 26148). The resulting purified and PCR-amplified DNA was subjected to standard quality control checks, including qPCR, Bioanalyzer analysis, and KAPA Library Quantification (Roche, KK4824). Sequencing was performed with unique single indexes on the Illumina NovaSeq 6000 Sequencing System, generating 200 bp reads.

### Hi-C pre-processing and interaction calling

Follows the pipeline as a recent study described (19). Paired-end reads from each replicate were pre-processed using the HICUP pipeline v0.7.4(13), aligned by bowtie2 with hg38 as the reference genome. The alignments files were parsed to *pairtools* v0.3.0 to process and *pairix* v0.3.7 to index and compress, then converted to Hi-C matrix binary format. *cool* by cooler v0.8.11 at multiple resolutions (500bp, 1, 2, 4, 10, 40, 500kbp and 1Mbp) and normalized with ICE method(20). The matrices from different replicates were merged at each resolution using *cooler*. Mustache v1.0.1(21) and Fit-Hi-C2 v2.0.7(22) were used to call significant intra-chromosomal interaction loops from merged replicates matrices at three resolutions 1kb, 2kb, and 4kb, with significance threshold at q-value < 0.1 and FDR < 1 × 10^−6^, respectively. The identified interaction loops were merged between both tools at each resolution. Lastly, interaction loops from all three resolutions were merged with preference for smaller resolution if overlapped.

### Definition of cis-Regulatory Elements (cREs)

We intersected ATAC-seq open chromatin regions (OCRs) of each cell type with chromatin conformation capture data determined by Hi-C/Capture-C of the same cell type, and with promoters (−1,500/+500bp of TSS) defined by GENCODE v40.

### 16 autoimmune traits’ GWAS summary statistics reformatted

**Table S3**Error! Reference source not found. lists the studies and accessions from which we drew GWAS summary statistics for each trait. ***LDSC*** (https://github.com/bulik/ldsc) recommends reformatting the summary statistics files using *munge_sumstats.py*. The table shows the number of total variants reported for each trait from the original studies, and the numbers after being filtered by *munge_sumstat.py*. This step also produced basic metadata about the summary statistics, such as mean chi^2^, max chi^2^, lambda GC, and mean value of the signed summary statistic column (beta, odd ratio - OR). Due to the inability of ***LDSC*** to produce meaningful heritability *h^2^* for traits with low numbers of variants, we applied *--merge-alleles* with the list of HapMap3 variants that ***LDSC*** used to estimate LD Scores (https://data.broadinstitute.org/alkesgroup/LDSCORE/w_hm3.snplist.bz2). This standardized all the GWAS sumstats files to the 1,217,311 variants from HapMap, discarded all the unmatched, and imputed the missing variants into the sumstats with *NULL* beta value.

### Partitioned heritability for cell type specific annotations with each autoimmune trait

We applied S-LDSC (stratified linkage disequilibrium score regression) using ***LDSC v.1.0.1*** to quantify to contribution to SNP-based effect-sizes and heritability to 16 complex immune diseases from each cell type’s 5 defined sets of input genomic regions: (1) OCRs, (2) OCRs at gene promoters, (3) cREs, (4) cREs with an expanded window of ±500 bp and (5) OCRs that were not cREs nor overlapped with gene promoters. Each set of input regions from each cell type was used to create the annotation, which in turn was used to compute annotation-specific LD scores for each cell types regions of interest set. These annotation-specific LD scores were used with 63 categories of the full baseline model. (v2.2) as control. To ensure comparability between traits from different studies, we used *--samp-prev* and *--pop-prev* to flag the sample and population prevalence for each trait, so that ***LDSC*** estimates SNP-heritability on the liability scale. The number of cases and controls were used to compute sample prevalence of each trait. We used the most recently reported trait prevalence for the European population for each trait, noted in **Table S3**. Two statistics we used to assess how effectively our annotations capture causal variation are heritability enrichment and standardized effect size (τ*), as previously defined(23). When conditioning two correlated annotations in a joint S-LDSC model, they may show similar enrichments, but the τ* for the annotation with a higher true causal variant membership will be larger and more positive.

### Genetic loci included in variant-to-genes mapping

For three autoimmune traits whose studies performed fine-mapping analysis, we used the significant variants within their provided credible sets. For the other traits, we leveraged their reported variants from associated loci which reached genome-wide significance within original studies. Proxies for each lead variant were queried using TopLD(24) and LDlinkR tool(25) with the GRCh38 Genome assembly, 1000 Genomes phase 3 v5 variant set, European population, and LD threshold of r^2^≥0.8 (results in **Table S3, column “N proxies”**).

### Differential analysis and clustering of correlated genes

Normalized transcripts per million (TPM) of all measured genes in 46 of 57 cell types was used to perform differential analysis using ***DEseq2*** package(26), where cell type and system (immune, metabolic, neural and other) were used as variables for the modeling contrast. Because many of the genes we gathered from the variant-to-gene mapping were lowly expressed in corresponding cell types or others, causing relatively high levels of variability, we used *apeglm* method for effect size (logarithmic fold change estimates) shrinkage(27) to alleviate this phenomenon during the genes ranking. Weighted correlation network analysis (***WGCNA***) package(28) was used to cluster genes from the variant-to-genes mapping process. WGCNA network construction power was chosen based on the analysis of scale-free topology for soft-thresholding. *blockwiseModules* with a power of 10 was used to create the correlation network and cluster genes into modules of similarly expressed genes.

### Pathways enrichment analyses

We performed two types of analyses on the set of genes from the variant-to-genes mapping process:

***Gene set over-representation analysis (ORA)*** was performed using ***clusterProfiler*** package(29) to identify GO biology process terms (org.Hs.eg.db databese) enriched within our genes set in each cell type. A relaxed P-value cutoff was set at 0.1, and the minimum including genes was set at 2 to ensure the capture of all possible enriched terms. An adjusted P-value of 0.05 was later used to filter the significant terms.

### Active-subnetwork-oriented gene set enrichment analysis

We used the ***pathfindR*** package(30) to identify active subnetworks in protein-protein interaction networks from Biogrid, KEGG, STRING, GeneMania, and IntAct databases, using the list of genes from the variant-to-genes mapping process. Then we provided the statistics from the differential expression analysis for ***pathfindR***, fold-changes were computed for cell type versus mean of other cell types, with cell system as a variable. We performed enrichment analyses on the identified subnetworks, discovering enriched KEGG pathways and GO biology process terms.

### Prediction of variant’s effect on transcription factor binding

Genomic positions (0-based coordinates) and allele alternatives of each proxy (from ***SNPlocs.Hsapiens.dbSNP155.GRCh38*** package with matching reference sequence from ***BSgenome*** package) were used to scan all position frequency matrix databases (from ***MotifDb*** package) for potential transcription factor binding disruptive effects. The motifbreakR function from ***motifBreakR*** package was used, with *filterp=TRUE* and setting a p-value *threshold=0.0005*, information content methods *method=’ic’* with even background probabilities of the four nucleotides *bkg = c(A=0.25, C=0.25, G=0.25, T=0.25)* and *BPPARAM = BiocParallel::SerialParam()* to allow serial evaluation.

### GWAS-eQTL colocalization

We used the same summary statistics for autoimmune disorders as in the ***LDSC*** analysis. We extended the proxies set further to include all available populations from the references (“EUR, AFR, SAS and EAS for TopLD and “ALL” for LDlink). For all the lead SNPs that had proxies identified in variant-to-gene mapping analysis on immune cell types, their chromosome coordinates were extracted, further augmented and consolidated into regions set with three key intervals: block positions, proxy-associated positional ranges, and a ±250 kb extended window. Using *GenomicRanges::reduce*, overlapping intervals were merged into a minimal set of non-redundant regions. For regions with multiple overlapping leads, pairwise linkage disequilibrium (LD) was assessed using the *LDmatrix* function from the LDlink API, and correlation coefficients (r^2^) were calculated. Regions with high LD (r^2^ ≥ 0.8) were grouped and merged, whereas those with lower LD (r^2^ < 0.8) were separated and further reduced to create 258 non-overlapped loci. We used eQTLs from 4 databases: eQTL Catalogue(31) (80-190 donors, available at https://github.com/eQTL-Catalogue), the Database of Immune Cell EQTLs (DICE eQTL(32), 91 donors, available at https://dice-database.org in GRCh37 reference genome, lifted to GRCh38 using UCSC LiftOver tool and LiftRsNumber.py), and the OneK1K database(33) (982 donors, available at https://onek1k.org). We overlapped these 258 immune GWAS loci with the significant gene expression eQTLs associations (eGenes, FDR > 0.05) for 18 cell type-specific data sets from 5 studies (QTS000002, QTS000003, QTS000012, QTS000024, QTS000026) of eQTL Catalogue, the 12 sets (B_CELL_NAIVE, CD4_NAIVE, CD4_STIM, M2, MONOCYTES, NK, TFH, TH1, TH2, TH17, TREG_NAIVE, TREG_MEM) of DICE, 9 sets (bin, bmem, plasma, cd4nc, cd4et, monoc, mononc, nk, dc) of OneK1K that correspondent to our library of immune cell types (details in **Table S4**) to narrow the list of genes that are regulated in response to immune diseases associated variants. For each gene implicated by our variant-to-gene pipeline, we extract available eQTL summary statistics within the overlapping locus: association P values, effect size estimate (β), and standard error of β; minor allele frequencies were included for OneK1K and eQTL Catalogue, and inferred from dnSNP155 for DICE. To assess the evidence for colocalization of shared causal variants for each gene between eQTLs of each cell types and all GWAS of each disease included in each overlapping locus, we first performed Approximate Bayes Factor colocalisation analysis under a single causal variant assumption using the *coloc.abf()* function. Subsequently, to account for the possibility of multiple causal variants, we adopted the Sum of Single Effects (SuSiE) framework for fine mapping using the *coloc.susie()* function, both implemented in the R package ***coloc***(34) and *susie_rss* from ***susieR***(35).

### Single-cell RNA-seq analysis

We collected immune single-cell RNA-seq for the autoimmune diseases, focused on the data with healthy control and no comorbidity and prioritized processed data from peripheral blood samples and with cell identity labels. The collected datasets are in **Table S3**. Origin tissues from which the immune cell types differentiated from were indicated in the table. We applied differential gene expression analysis comparing disease versus control on each separate cell type population of each data set, using the default Wilcoxon Rank Sum test of the ***Seurat*** package(36); p-values were FDR adjusted.

## RESULTS

### Large-scale survey of 3D cis-regulatory architectures identifies target tissues enriched for autoimmune disease loci

Genes are frequently controlled by regulatory elements that are far away in the linear sequence of the genome, but due to the highly folded structure of chromatin in the nucleus, they come into close proximity in three-dimensional space. The importance of this long-range mode of gene regulation was first appreciated at the *Hbb* and *IFNg* loci(37–39), and has been confirmed at a whole genome scale by studies showing that the set of genes contacted by distant regulatory regions exhibit 10-to 20-fold higher levels of expression than genes not in contact with distant cRE(40). Therefore, a spatial map of genome structure in a given cell type represents a crucial tool for understanding any gene’s full *cis*-regulatory architecture and how genetic variants might contribute to its expression.

We used ATAC-seq and the chromosome conformation capture technologies of promoter Capture-C (PCC) and Hi-C to map the promoter-connected open chromatin (putative cRE) landscapes of 57 human cell types representing immune, metabolic, neuronal, and pluripotent stem cell lineages (**Table S1**). All datasets were generated by our laboratory using our standardized wet- and dry-bench pipeline and include 9 immune cell types not previously published (EBV-B, T helper 1, T helper 2, T helper 17, T regulatory 1, T regulatory, plasmacytoid dendritic, CD1c+ dendritic, and natural killer cells).

To determine which of these tissues may be most impacted by autoimmune disease-associated genetic variation, we measured the heritability enrichment (**Figure S1A**) and standardized annotation effect size (τ*, **Figure 1A,Figure S1B**) across the 3D cRE landscapes of each cell type by stratified linkage disequilibrium score regression (S-LDSR)(41) using recent European-ancestry GWAS summary statistics of sixteen autoimmune diseases: allergy (ALG), eczema (ECZ), ankylosing spondylitis (AS), psoriasis (PSO), Crohn’s disease (CRO), ulcerative colitis (UC), celiac disease (CEL), inflammatory bowel disease (IBD), rheumatoid arthritis (RA), juvenile idiopathic arthritis (JIA), multiple sclerosis (MS), systemic lupus erythematosus (SLE), type I diabetes mellitus (T1D), Hashimoto’s thyroiditis (HT), Graves’ disease (GD) and vitiligo (VIT) (**Table S3**). Genetic heritability for the risk of the majority of autoimmune diseases showed statistically significant enrichment with positive effect sizes in the cRE landscapes of most or all of the 22 analyzed immune cell types and states, and was largely not enriched in metabolic, neuronal, and pluripotent cell types (full detail in **Table S5**).

**Figure 1.**
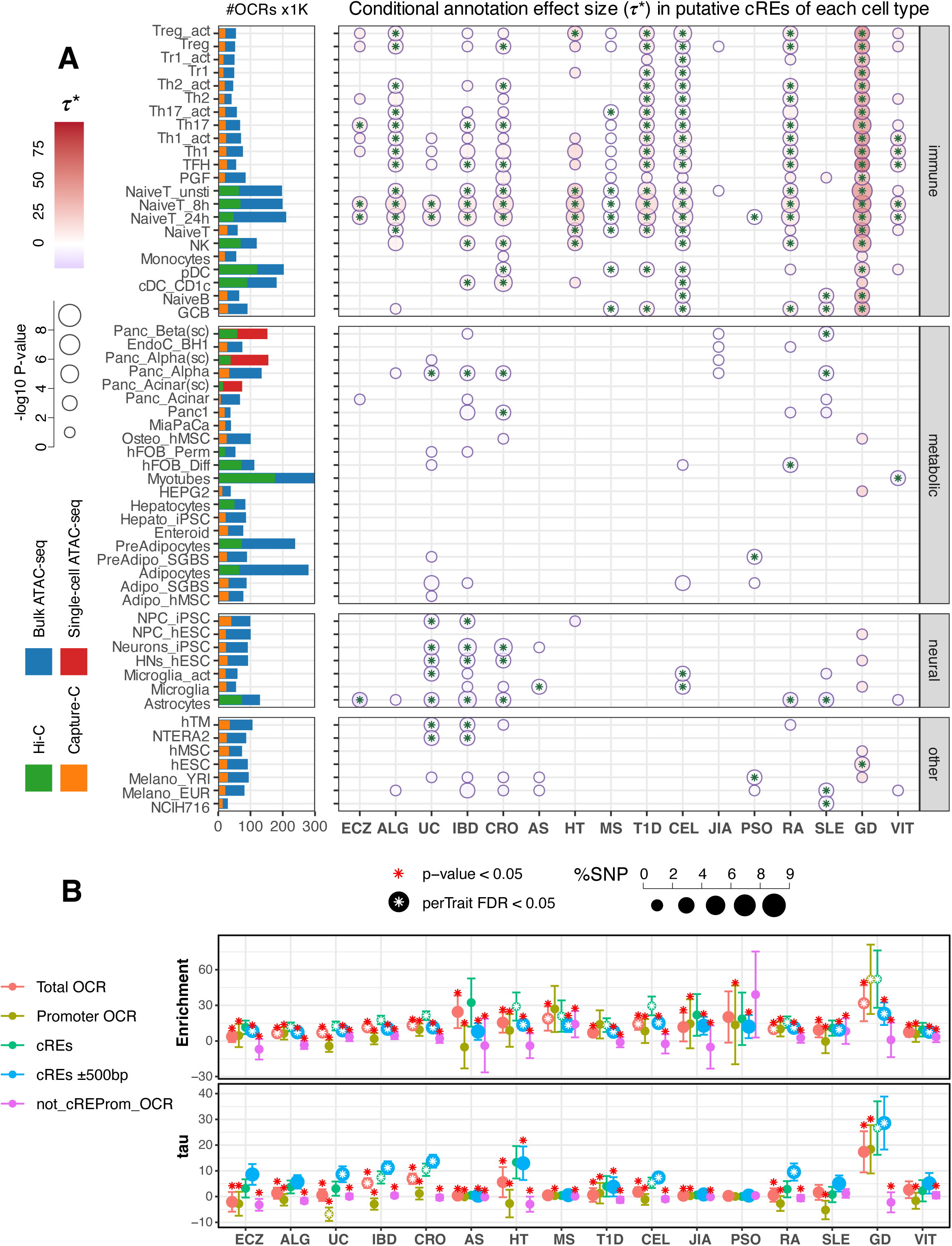
Autoimmune disease conditional effect size (t*) within putative cREs annotation across diverse cell types. **A**. Bar plots (left) depict total number of open chromatin regions (OCR x1000) identified by bulk (blue) or single-cell (red) ATAC-seq and the proportion of those OCR contacting a gene promoter (putative cRE) as measured by HiC (green) or promoter capture-C (orange). The dot plots depict significant (p-value ≤ 0.05) conditional effect sizes within cREs annotation for each cell type across 16 autoimmune traits as determined by S-LDSC analysis. The colors of the dots correspond to τ* values, with dots featuring an asterisk indicating FDR ≤ 0.05. The size of the dots corresponds to p-values in - log10. **B**. 2 dot plots depicting heritability enrichment (top) and conditional effect sizes τ* (bottom) for CD1c+ DC cells for each immune trait, grouped by annotations: all OCR (red), promoter OCR (light green), promoter-contacting OCR (cRE, green), cRE expanded to 500 bp flanking regions (blue), and OCR not contacting/overlapped a promoter (violet). Red asterisks above indicate a significant p-value<0.05 in one-side (lesser) T-test comparing other categories with the green putative cRE. White asterisks inside dots indicate significant p-values ≤0.05 from LDSC analysis.

Our cRE annotations could not capture heritability for JIA, AS and PSO, as these traits were associated with the fewest immune cell types showing either aggregated enrichment or high τ*. Grave’s disease displayed very strong enrichment for the most cell types, including for immune, metabolic, neural and tumor (**Figure S2**). Low global GD heritability (h^2^=0.0061) likely contributes to the high adjusted contributions of cREs to per-SNP heritability in this case. Similarly, for traits with lower global heritability such as ALG (h^2^=0.055), HT (h^2^=0.01) and T1D (h^2^= 0.0389), high impact variants (relative to the global level) were captured more than the baseline annotations by immune cell cREs and attributed to the total enrichment. The opposite direction was observed for ECZ (h^2^=0.0457), UC (h^2^=0.2831), and SLE (h^2^=0.4244). These traits exhibited highly significant enrichment, indicating that immune cell cREs captured a substantial fraction of heritability relative to their genomic span. However, this was accompanied by low or insignificant τ* values (**Figures 1, S3**), suggesting that the contribution either 1) overlaps substantially with other baseline annotations in the model, diluting its independent effect, 2) is localized and driven by a small subset of SNPs within cREs, and/or 3) reflects the relatively smaller per-SNP contributions due to the high global h^2^. Higher heritable traits such as CEL (h^2^=0.3421) maintained significance from aggregated enrichment to conditional per-SNP effect sizes captured by immune cREs.

Statistically significant enrichment (*P*<0.05) instances were observed in all 22 immune cell types for at least one immune trait (**Figure S3**). The cRE landscapes of naïve and activated CD4+ helper T cells exhibited enrichment for 15 out of 16 immune traits and captured significantly high τ* for the majority, while naïve T cells cRE singularly capturing the effects of JIA and PSO heritability. Monocytes showed enrichment for only 7 traits and significant τ* values only with 2 traits, namely CRO and GD. The regulatory landscapes of naïve and GC B cells and EBV-transformed PGF B cell line were most highly enriched for SLE heritability and demonstrated strong enrichment for RA and CEL, consistent with the clear role of autoantibodies in driving these conditions^44^. Conversely, ECZ, AS, JIA, and PSO variants were not enriched in B cells. Differentiated effector T cell (Th1, Th2, Th17) landscapes were highly enriched for most autoimmune traits, but not for AS, JIA, PSO and non-significant τ* values for SLE. Th17 regulatory landscapes were particularly enriched in ECZ and MS, diseases known to be driven by Th17-derived cytokines(42–44). TR1 and TFH cells exhibited moderate heritability enrichment across many diseases, with TFH landscapes exhibiting particularly strong enrichment in RA heritability. NK regulatory architectures were most significantly enriched for UC, CRO, and IBD heritability but higher τ* for HT and GD.

The cRE landscapes of monocytes, CD1c+ DC (cDC2), and interferon-producing pDC cells were enriched for heritability for multiple diseases, but were not enriched for ECZ-or PSO-associated variants, and deficient in capturing effective variants for ALG, UC, AS, JIA and SLE. Treg cells stood out as the most highly enriched cell type for T1D GWAS loci, and featured prominently in VIT, RA, HT, ECZ, UC, CRO, and IBD heritability. This highlights the broad involvement of autoimmune susceptibility variants, specifically in regulating gene expression within Treg cRE regions that do not overlap with other regulatory annotations. These findings align with established role of Treg cells in self-tolerance and regulation of inflammation(45,46). B cell types were the only immune cells exhibiting highly significant SLE heritability and effect size(47,48) within their cRE landscapes. Interestingly, the incidence and pathobiology of these diseases have been linked to EBV infection(49–52), suggesting possible genetic interactions between EBV and autoimmune variants in infected B cells. These findings underscore the diverse and complex interplay of immune cell types in autoimmune disease susceptibility.

The 3D regulatory architectures of some non-immune cell types also exhibited enrichment for several immune traits (**Figure S1**, **Figure S3, Table S5**). For instance, adipocyte lineage cells showed significant enrichment for ECZ, ALG, HT and RA heritability, and enrichment for ALG and ECZ heritability was found on the adipocyte/osteoblast axis, including hFOB (fetal osteoblasts), hMSC-derived osteoblasts, pre-adipocytes, adipocytes, hMSC-derived adipocytes, hepatocytes and colon-derived enteroids. Significantly, neural progenitor cell cRE landscapes were enriched in heritability for MS, an autoimmune disease targeting nervous tissue in the CNS, and the cRE landscape of intestinal organoids (enteroids) was strongly enriched for UC heritability. However, the conditioned effect sizes captured by cREs of these enriched non-immune cell types were low to negative, suggesting the average effect per SNP is small within these cREs. GD heritability was enriched in the greatest number of non-immune cell types, including Panc1, MiaPaCa (pancreatic ductal cancer cell lines), osteoblast (hMSC-derived), HEPG2, NPC-hESC, hypothalamic neurons (hES-derived), microglia, activated microglia, hMSC, hESC, melanocyte-YRI, NCIH716 (colorectal adenocarcinoma). But only a few cell types cREs retained significant but low effect sizes τ* after conditioned with baseline annotations, suggesting these enrichments were not unique for cRE within these cell types. Notably, while UC, CRO, and IBD were not significantly enriched within neural cell types, the conditional effect sizes associated with neuronal cREs were significantly negative. This finding suggests a potential protective role for variants captured within neuronal and glial cRE, aligning with evidence that certain neural cell types, particularly enteric glial cells, contribute to protection in inflammatory bowel diseases(53). For instance, alterations in neurotransmitter expression within the myenteric plexus have been observed in IBD(54), and colitis has been shown to induce rapid enteric neurogenesis as a compensatory mechanism to maintain gut function during inflammation(55). These observations suggest that some UC-associated variants may exert their effects through non-immune intestinal cells, further highlighting the protective influence of neuronal and glial elements in modulating disease risk(56).

### 3D cRE architectures are enriched for autoimmune disease heritability compared to 1D epigenomic feature maps

Importantly, these LDSC analyses show that 3D promoter-contacted open chromatin (putative cRE) landscapes specifically and consistently exhibit the strongest heritability enrichment across all cell types compared to open chromatin landscapes not informed by Hi-C/PCC or compared to open chromatin proximal to gene promoters (**Figure 1B**). The magnitude of heritability enrichment for each immune trait using cell type cRE landscapes was significantly greater in 72% (2616 of 3648) of pairwise comparisons (red stars in **Figure 1B**). Moreover, the subset of open chromatin regions not detected as interacting with a gene promoter by Hi-C/PCC consistently exhibited the lowest level of heritability enrichment and generally failed to reach statistical significance compared to other biological constraints tested. The originally reported LDSR method extended the analysis to the ±500 bp regions flanking their regulatory element categories(41). Expanding our analysis similarly to include ±500 bp flanking our defined cRE resulted in the inclusion of more weighted variants (represented by larger blue dots in all panels in **Figure 1B**), more enriched cell types (**Figure S2B**, **Figure S3B**), decreased enrichment range across cell types, decreased 95% CI, and decreased level of significance. Interestingly, we observed a striking contrast: promoter OCRs within immune cell types captured significant effect size variants from far fewer traits compared to other OCR annotations, whereas promoter OCRs in non-immune cell types accounted for the majority of trait heritability. However, the majority of the heritability that promoter OCRs of non-immune cell types captured were negative effect and across most of the traits (**Figure S3C,D**). These comparative metrics underscore the enhanced predictive value of 3D chromatin-based maps of putative gene regulatory architecture for assessing the heritability of autoimmune traits across tissue types.

### Chromatin-based V2G maps reveal trait-specific genetic influence on disease risk via genes shared across traits and cell types

Having observed that GWAS-informed open chromatin-promoter interactomes are highly enriched for autoimmune heritability in multiple immune cell types, we elected to next determine which genes are in contact with, and therefore likely regulated by, accessible autoimmune variants by systematically mapping the genomic positions of linkage disequilibrium (LD) proxies within the cRE landscape of each cell type. This approach cannot confidently identify co-inherited proxies in the highly polymorphic and extended LD landscape of the MHC region on chromosome 6, so in line with other variant-to-gene efforts we excluded this chromosomal region from our analysis. These 3D autoimmune variant-gene mappings revealed 5,488 genes targeted by chromatin contacts with 3,642 proxies connected to all immune traits in all cell types (full result in **Table S6**). While most proxy variants were unique to a GWAS sentinel in a given cell setting, some proxies were consistently observed across different immune traits (**Figure 2A**). This allowed us to categorize target genes as unique vs. shared based on their contacts with trait-specific vs. shared variants. This led to a combinatorial complexity of over 25,000 variant-gene pairs (**Figure 2B**), and **Figure 2C** depicts examples of two such combinations. Most of the implicated variants were trait-specific, with only 18% (679 proxies) shared between traits (**Figure 2D**, detailed in **Figure S4A**). Notable outliers were rs34536443, rs35350651 and rs653178 on chromosomes 12 and 19 that were consistently observed with seven autoimmune traits. On average, a single variant contacted 4 gene promoters in a specific cell type through chromatin loops. Immune cells exhibited a statistically significantly higher number of autoimmune variant-gene contacts, and thus higher total number of implicated genes, compared to non-immune cell types (**Figure S4B**), with pDC, cDC2, and NK cells exhibiting significantly more variant-gene contacts than other immune cell types.

**Figure 2.**
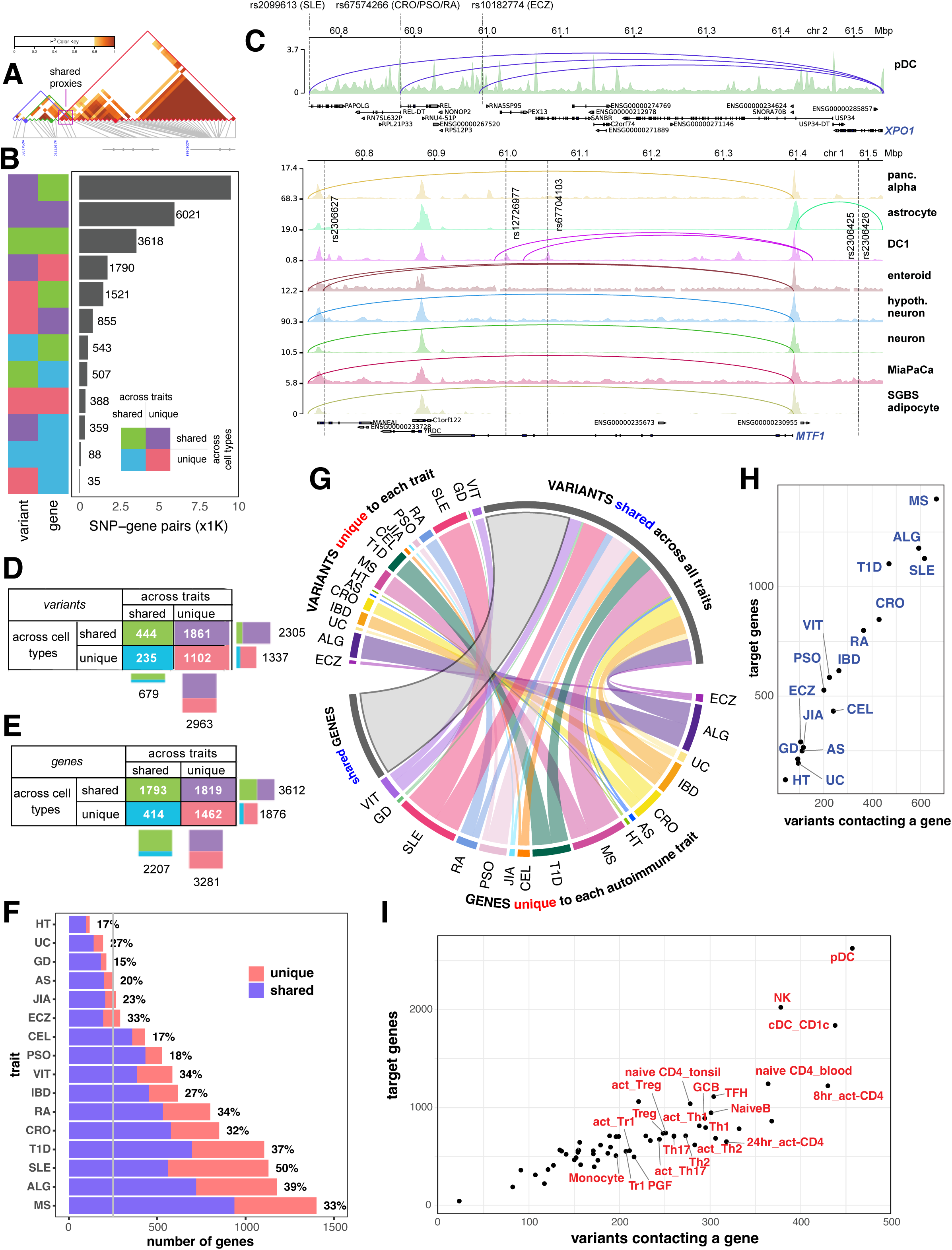
3D chromatin-based variant-to-gene mapping across autoimmune diseases and cell types. **A**. Schematic shows 3 lead SNPs from 3 different traits can share proxies. **B**. Bar plot depicting the number of variant-gene pairs in which the variant (left column) or the gene (right column) is unique vs. shared across traits and cell types (green - shared across traits and cell types, purple - unique to trait but shared across cell types, cyan - unique to cell type but shared across traits, red – unique to trait and cell type). **C**. Genomic views of *XPO1* gene locus, contacted by 3 variants from 5 different traits uniquely detected in pDC (top), and of the *MTF1* gene locus, contacted by 5 RA variants but detected in 7 different cell types (bottom). **D**. Detail of variants unique vs. shared across traits and cell types. **E**. Detail of genes unique vs. shared across traits and cell types. **F**. Bar plot shows numbers of V2G genes of each trait, colored by whether they’re uniquely identified for that trait (percentage number) or shared with other traits. **G**. Chord diagram shows proportions of genes (g.) and variant (v.) of each trait are trait-specific (color-coded) or shared (grey). **H**. Scatter-plot depicting target gene number versus variant number for each trait. **I**. Scatter-plot depicting target gene number versus variant number for each cell type with immune cell types indicated.

While implicated variants were largely trait-specific, 40% of target genes of these variants (2,207 of 5,488) were implicated across more than one autoimmune disorder (**Figure 2E**, detail in **Figure S5A**), with 11 genes implicated across a maximum of 9 traits (*e.g*., *TYK2, ICAM1, ICAM3, TNFIP3, CLEC16A, BACH2*). Genes implicated in B cells were generally widely shared among other immune cell types (**Figure S5B**), while T cell subsets and dendritic cell types showed the most exclusive pattern of V2G-implicated genes. Among the traits, SLE had the highest number of exclusively implicated genes (569), followed by ALG (456) and MS (465) (**Figure 2F**).

The lower degree of exclusivity for target genes compared to that of variants across traits is due to the fact that nearly half of the trait-specific variants end up targeting the same genes (**Figure 2G**). An example is the *XPO1* gene, whose promoter was contacted by multiple variants from various immune diseases, but only in pDC (**Figure 2B**). The degree of overlap of implicated variants and target genes across cell types was much higher than across traits, with 63% of variants (2,305 out of 3,642 **Figure 2D** and **Figure S4A**) and 66% of genes (3,612 out of 5,488, **Figure 2E** and **Figure S4B**) implicated across two or more cell types. Only 388 variant-gene pairs were unique to both trait and cell type.

The total number of variants vs. target genes implicated per trait (**Figure 2H**) or per cell type (**Figure 2I**) were highly concordant. Overall, these analyses show that the majority of variant-gene pairs are composed of target genes shared across traits and cell types and implicated variants that are largely trait-unique but shared across multiple cell types. These results suggest that putatively functional variants that are uniquely observed with single autoimmune traits often converge on the same genes across multiple cell types and therefore may ultimately share common mechanisms of influence on disease risk, potentially via haplotypic effects.

### Chromatin-based V2G maps implicate both common and unique biological pathways targeted by autoimmune-associated genetic variation

To explore how complex patterns of 3D autoimmune variant-gene contacts across traits and cell types may drive defined biological pathways to influence disease risk, we conducted pathway analysis for genes implicated in each cell type for each trait. This analysis focused on pathways supported by a minimum of five implicated genes and enrichment was weighted by protein-to-protein interaction subnetworks and by the degree to which pathway genes show higher or lower expression in a given cell type compared to their average expression across all cell types. This analysis yielded 2,240 significantly enriched GO terms and 310 KEGG pathways (**Table S7** and **Table S8**). KEGG pathways enriched across all or a majority of autoimmune traits were cytokine-cytokine receptor interactions, bacterial infection (*Salmonella, Yersinia*), viral infection (HTLV-1, Herpes, COVID-19 disease), NFkB and MAPK signaling, C-type lectin and Toll-like receptor signaling; and top shared GO terms were transcription factor binding and ubiquitin protein ligase binding. These processes have experimentally- and clinically established roles in innate and adaptive immunity, immune tolerance, and autoimmunity, further validating the biological relevance of the 3D cRE architectures reported here.

The 66 genes within the cytokine-cytokine receptor interaction pathway were enriched in 21 immune, 14 metabolic, and 3 neural cell types (**Figure S6**), 36 genes involved in the *Salmonella* infection pathway were enriched in 21 immune, 11 metabolic, and 6 neural cell types (**Figure S7**). Genes involved in DNA transcription factor activity were enriched in 18 immune, 8 metabolic, and 5 neural cell types, and the ubiquitin ligase pathway was enriched across 21 immune, 10 metabolic, and 5 neural cell types. Colon-derived enteroids from IBD patients(57) offer an interesting example: the 3D regulatory architectures of these intestinal organoids showed enrichment for UC heritability at genome-scale (**Figure 1A**), and a large number of genes (>700) in these cell types were connected to UC, IBD and CRO variants. However, the implicated genes are not all involved in the same biological processes, as the pathway analysis shows that only one-third of the pathways enriched for all three traits in these organoids are shared between CRO and UC, while another third only enriched in IBD, and the final third specific to UC (**Figure S8**).

While most of the enriched pathways are shared across traits, 31 KEGG pathways show trait-specific enrichment (**Figure S9**), and many of these point to processes known to influence disease pathology. For example, the ‘maturity onset of diabetes in the young’ pathway is enriched in our T1D gene set, but as might be expected, not in any other gene sets implicated by other autoimmune traits. Nicotinamide and selenium levels are associated with SLE pathology and as supplements have been shown to ameliorate symptoms in patients(58–62), and both ‘nicotinamide metabolism’ and ‘seleno compound metabolism’ were uniquely enriched in the SLE-implicated gene set. One pathway uniquely enriched in the MS-implicated gene set is ‘pyrimidine metabolism’, and levels of pyrimidine metabolites in cerebral-spinal fluid is associated with symptom severity in MS patients, and therapeutic inhibition of pyrimidine biosynthesis ameliorates disease(63,64). The only pathway specifically enriched in JIA is ‘nicotine addiction’, and interestingly, maternal smoking and second-hand smoke exposure is a known significant risk factor in JIA(65–67).

### Comparison of 3D chromatin-based vs. eQTL-based effector gene nomination approaches

To determine whether our GWAS-implicated variant-gene contacts correlate with variant-gene quantitative trait effects, we compared the set of genes implicated for each trait in each cell type reported in this study with the set of eGenes identified in eQTL studies in the same or similar cell/tissue types from GTEx whole blood, DICE, OneK1K and eQTL_Catalogue datasets. The genetic locations of all proxies in high-LD with lead variants from our chromatin-based V2G analysis were clustered to construct 285 loci, and used to extract eGenes from each database whose expression was significantly associated with at least one of the variants within these loci (**Methods**). This yielded 11,182 eGenes from whole blood GTEx, 14,168 eGenes from DICE, 3,945 from OneK1K, and 8,033 from eQTL_Catalogue datasets across various immune cell types, compared to 4,125 genes implicated within immune cell types by our 3D chromatin-based approach (**Figure S10**). Twelve loci yielded no eGene from any eQTL datasets on chromosomes 5, 6, 8, 9 and 21 (**Figure S11**, **Table S9**).

To compare V2G genes and eQTL eGenes on an equal basis, we then focused exclusively on eGenes linked to cis-eQTLs with high-LD variants and located on the same chromosome as the locus (**Figure 3A**, **Figure S11**). GTEx and 3D chromatin V2G implicated 1,265 locus-specific eGenes (1276 pooled loci) in common across all 22 immune cell types and 16 disease traits, meaning that our 3D chromatin V2G implicated 20% (from 6276) of the GTEx whole blood eQTL within these loci, while the GTEx whole blood eQTL dataset detected 31% of the genes implicated in immune cells by our 3D chromatin V2G approach. The cell type-specific eQTL from all datasets and our 3D chromatin V2G implicated 779 locus-specific eGenes (2139 pooled loci) in common if disregarding the corresponding cell type, so that only 4.3% of the (total 18042) eGenes implicated within these 258 loci from all cell type-specific eQTL datasets were evidenced by 3D chromatin V2G, while 36% of the 3D chromatin V2G approach was in concordant with the cell type-specific eQTL datasets.

**Figure 3.**
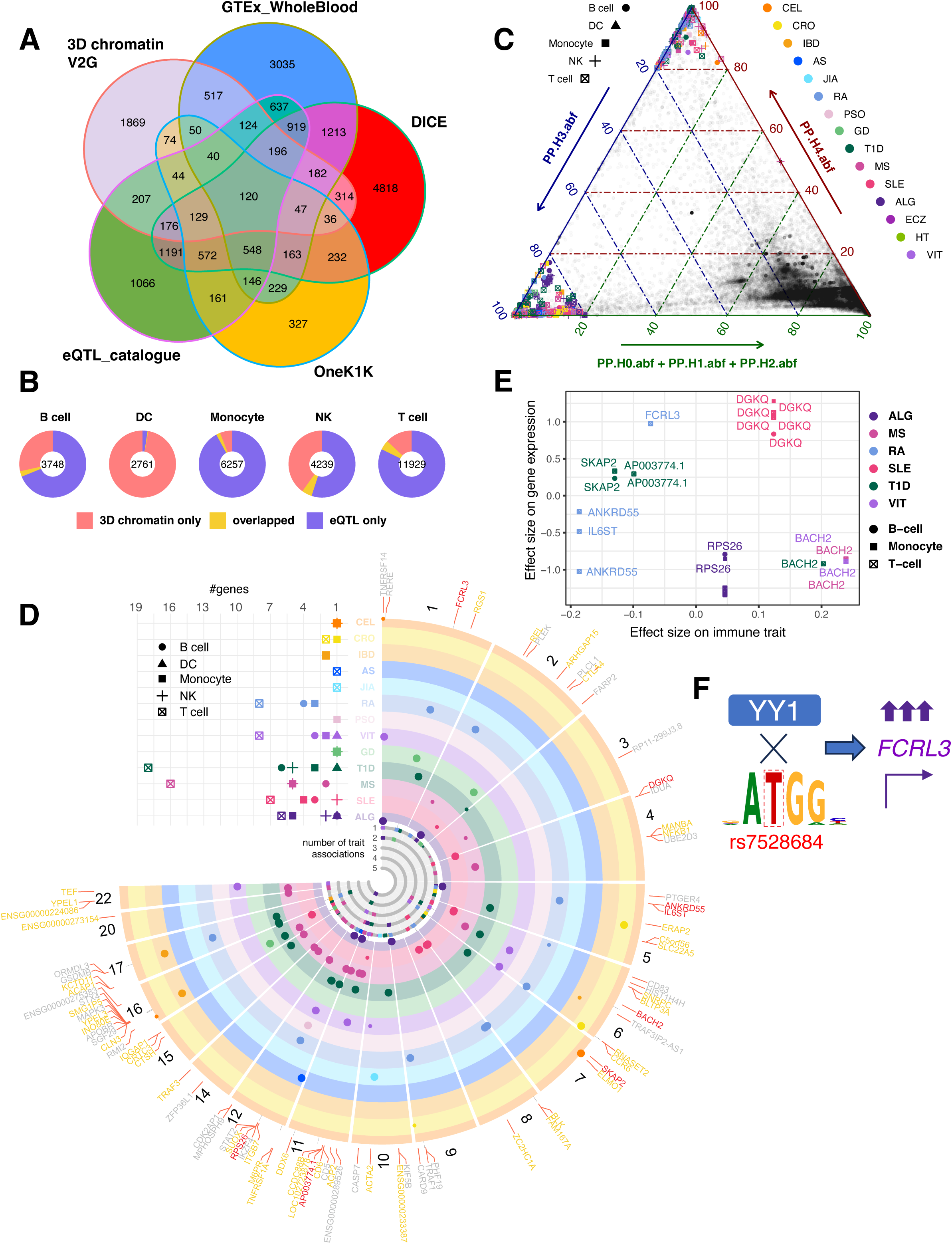
Comparison of eGenes implicated by 3D chromatin-based V2G vs. eQTL. **A**. Shared eGenes across different eQTL datasets. **B.** Pie charts of eGenes identified by eQTL that match (light orange) or do not match (purple) the gene identified by 3D chromatin V2G (grey) for each cell groups, all traits collapsed. **C.** Ternary plot shows results from coloc.abf analysis, those with HH.P4.abf≥0.8 were highlighted on top corner and HH.P3.abf≥0.8 were highlighted in left corner**. D.** Fuji-plot presents the genomic distribution of immune disease loci exhibiting high-confidence colocalization (PP.H4.abf ≥ 0.8 from panel C); tracks display loci colored by associated immune trait, with dot size proportional to the number of associated traits (pleiotropy); central panel shows the count of associated traits per gene, stacked and colored by trait; top quarter dot plot details the number of colocalized genes per trait track, stratified by cell type groups; gene names are colored to reflect causal variant confidence: light orange for coloc.abf-identified high-confidence variants (SNP.PP.H4.abf ≥ 0.8), grey for no high-confidence coloc.abf variant, and red for V2G approach matches. **E**. The dot plot visualizes the relationship between variant effect sizes on eGene expression (x-axis) and immune trait risk (y-axis) for 8 matched variants from panel D; colors denote the specific immune trait, and shapes indicate the cell type group in which the eQTL was identified. **F**. Variant rs7528684 disrupts YY1 motif binding, potentially upregulating *FCRL3*.

Mapping eGenes from the matching cell types from eQTL datasets and the corresponding immune diseases from V2G dataset reduced the number of overlapped events significantly. Overall, the overlap between these two gene nomination strategies remains relatively limited, while also dependent on the availability of the eQTL for corresponding cell types (**Figure 3B**, detailed in **Figure S12**). MS yielded the highest number of commonly implicated eGenes across all cell types (278), followed by ALG (260), SLE (229) and T1D (216), and naïve CD4+ T cells yielded the highest number of commonly implicated eGenes across all autoimmune traits (347), followed by NK cells (222) in concordant with the abundance of their V2G genes, while dendritic cells yielded least number of common eGenes despite having the highest V2G genes. In proportion to the total genes implicated by 3D chromatin V2G, HT had the most evidenced by eGenes (45%), followed by JIA (42%), AS (38%), while naïve CD4+ T cells (21%) monocytes (21%) and regulatory T cells (13%) leads on the cell types scales (**Table S10**). These findings suggest that 3D chromatin-based V2G approaches are more effective at identifying eQTL-based eGenes compared to the ability of eQTL methods to identify 3D chromatin-based target genes.

We used the approximate Bayes factor (*abf*) framework of *COLOC*(34) and the iterative Bayesian step-wise selection regression of *SuSiE*(35) to investigate the colocalization between each autoimmune trait with each cell type eQTL from each resource for each identified eGenes. Under the single-causal assumption of *COLOC* analysis, 81 genes within 55 loci showed a high estimated posterior probability of both disease and eQTL sharing a single causal variant (PP.H4.abf ≥0.8, **Figure 3C, Table S11**); 35 of these loci V2G approach identified more than one putative causal variants. For 8 of these 81 genes, the variant identified by V2G approach was found by *COLOC* in high confidence of being a common causal SNP shared (SNP.HH.P4≥0.8, red genes in **Figure 3D**). These genes were implicated in key immune processes: *RPS26* encodes a ribosomal protein required to protect activated T cells from p53-mediated apoptosis(68), *DGKQ* and *SKAP2* participate in T cell receptor and integrin signaling in immune cells(69), and a rare coding mutation in *SKAP2* has been linked to altered myeloid cell function in T1D(70). *FCRL3* encodes an inhibitory receptor on B cells and regulatory T cells(71,72), *IL6ST* encodes the *GP130* receptor for the pro-inflammatory cytokine IL-6, and *BACH2* is a central transcription factor that controls the differentiation of multiple T helper subsets including Treg, Th17, and TFH cells(73). Notably, the potential variants showed opposing effects on gene expression and immune disease risk for different genes. For *DGKQ, IL6ST* and *ANKRD55*, decreased expression correlated with reduced immune disease risk, and increased expression with higher risk. However, increased expression of *FCRL3*, *SKAP2*, and *AP003774* was associated with lower risk of T1D and RA, while decreased expression of *BACH2* and *RPS26* increased immune disease risk (**Figure 3E**). To explore potential mechanisms by which autoimmune-associated genetic variation may influence disease risk, we used *motifbreakR* to predict transcription factor binding site (TFBS) disruption by these high-probability causal variants. We found that these autoimmune risk variants are predicted to alter the affinity of NFkB, ETS, and forkhead family DNA binding (**Figure S13**). The T allele of rs7528684 in the promoter of *FCRL3* reported to be associated higher expression and confer RA protection (74),(75) was likewise predicted to lower the binding affinity of the transcriptional repressor YY1 (**Figure 3F**), potentially increasing *FCRL3* expression in naïve T cells. Since more than half (141) of the 258 loci tested harbored more than one potentially causal variant, we adopted the sum of single effects (SuSiE) framework for fine mapping in the presence of multiple possible causal variants with the posterior probability of both disease and eQTL being both associated high-LD variants within a locus (PP.H3.abf≥0.8). 110 loci met this threshold, and in 24 of which the variant identified by our V2G was in high confidence of being colocalized with other causal SNPs (**Table S12**).

### Functional validation of FDFT1 as a novel immunomodulatory drug target implicated by 3D chromatin-based V2G

Autoimmune GWAS represents the genetics of immune tolerance and immunoregulation; therefore, understanding the mechanisms by which autoimmune variants-target genes and biological pathways provide important disease insights. These could then be translated into novel immunomodulatory therapies tailored to specific autoimmune diseases or targeted at inflammation across multiple disorders including cancer and organ transplant rejection. The V2G maps reported here implicate a large array of genes and pathways in and across various immune cell types and autoimmune diseases. Many of these genes/pathways represent well-known biology, and some are currently the targets of FDA-approved therapies for immune-mediated disease. However, a large number of implicated genes, many of which are readily ‘druggable’, are novel in that they have not been studied in the context of immunology or assessed as an immunomodulatory target in an inflammatory context. Indeed, we have used this V2G approach in previous studies to nominate and functionally validate 12 novel immune targets with previously unappreciated roles in human T cell activation(76), follicular helper T cell function(77) and T cell help for B cell differentiation(40). We have also validated 3 novel non-immune targets that regulate bone mineralization(78,79), sleep behavior(80), and type 2 diabetes(81).

An example of a novel autoimmune drug target implicated in this study is FDFT1, a farnesyl-diphosphate farnesyltransferase (*aka*, squalene synthase) that catalyzes the first committed step of isoprenoid synthesis for cholesterol production. The *FDFT1* gene interacts with 7 MS-associated variants (**Figure 4A**, top panel), 7 SLE-associated variants (**Figure 4A**, bottom panel), and one variant associated with both RA and SLE (**Figure 4A**, middle panel). These variants are part of a diverse regulatory landscape at this locus, with SLE variants contacting *FDFT1* in naïve B cells, germinal center B cells, and microglia, and MS variants contacting *FDFT1* in pancreatic cells, iPSC-derived neural progenitor cells, TFH cells and Th17 cells (**Figure 4A**). Prior studies showed that *FDFT1* is downregulated in astrocytes in both mice and humans with multiple sclerosis(82,83), and neuronal induction of *FDFT1* and squalene synthase activity contributes to myelination in mouse models of chronic demyelinating disease(82), however, a role for *FDFT1* in the function of any of the immune cell types studied here has not been established.

**Figure 4.**
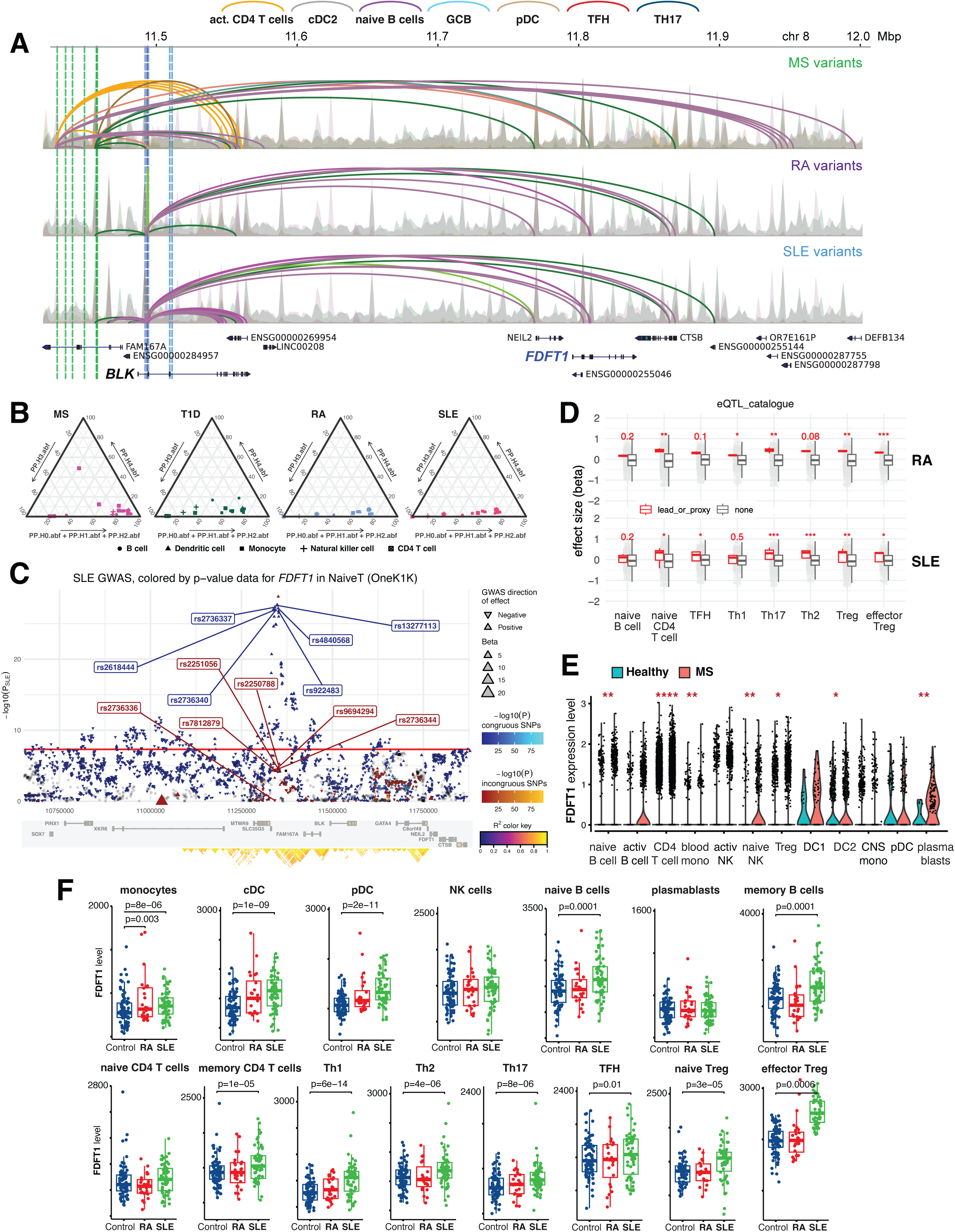
Functional validation of the 3D chromatin-based V2G target *FDFT1*. **A**. Genomic view at the *FAM167A-BLK* locus depicts loop contacts (arcs) between accessible proxies associated with MS (7, green vertical lines), SLE (7, grey vertical lines) and SLE+RA (1, blue vertical line) and gene promoters in activated CD4+ T cells (orange), cDC2 (grey), naïve B cells (purple), germinal center B cells (light blue), pDC (brown), TFH cells (red), and Th17 cells (dark blue). MS variants contact *FDFT1* in TFH and Th17 cells (top panels) and SLE/RA variants contact *FDFT1* in naïve B cells, germinal center B cells, and pDC (bottom panels). **B**. Ternary Diagrams of colocalization results within locus chr8:9921161-11972641 with eQTL of *FDFT1* across all cell type-specific datasets. **C**. eQTpLot for *FDFT1* in naïve T cells versus SLE shows chr8:9921161-11972641 locus for SLE GWAS, leads from V2G analysis were labeled, congruous effects are plotted using a blue color scale, in contrast to variants having an incongruous effect (in red), the size of each triangle is proportional to the eQTL normalized effect size (beta) in T cells, while the directionality of each triangle corresponds to the direction of effect of the variant on the GWAS trait. **D.** Boxplots comparing the effect sizes of various variants on the expression of *FDFT1*, grouped by cell type and immune trait; variants identified through the V2G approach are indicated by red boxplots; statistical significance between variant groups was assessed using a T-test, and the resulting p-values are shown. **E**. Differential scRNA-seq^65^ analysis of *FDFT1* expression in immune cell clusters from healthy donors (blue) or MS patients (red). *P<0.01, **P<0.001, ***P<0.0001, ****P<0.00001. **F**. Differential bulk RNA-seq^66^ analysis of *FDFT1* expression in sorted immune cells from healthy donors (blue), RA patients (red), or SLE patients (green). P values are shown.

To determine whether *FDFT1* expression is linked to autoimmune disease-associated genetic variation, we determined the distribution of posterior probabilities from colocalization analyses within the chr8:9921161-11972641 locus between each immune disease and eQTL of *FDFT1* across different cell types (**Figure 4B, Table S13**). SLE, RA, MS and T1D displayed high PP.H3.abf, consistent with our observation of SLE-, RA- and MS-associated proxies physically contact *FDFT1* through chromatin loops. There was high enrichment of concordant *FDFT1* eQTL signals from naïve CD4+ T cells among significant SLE variants (OneK1K, *P-value* ≤ 5e-08, **Figure 4C**), as well as in other differentiated T cell subsets, but unlike V2G, no B cell type was implicated in this analysis. We observed similar enrichment for RA signals, suggesting a significant association of this locus with the functions of immune T cells. The relatively low signal for MS within this locus resulted in no enrichment of any corresponding *FDFT1* eQTL (**Table S13**). Interestingly, although not as significant as SLE or RA, T1D variants with *P-value* ≤ 5e-06 within this locus were highly enriched with *FDFT1* eQTL in various T and B cells, albeit not implicated by 3D chromatin V2G (**Table S13**). For most combinations, this colocalization analysis supports the overlap of significant enrichment between the traits and eQTL at this locus but indicates separate signals instead of a single causal variant. Most of the putative variants we identified with our V2G approach significantly increase *FDFT1* expression in various classes of T cells and B cells (**Figure 4D**, **Figure S14**).

To determine whether autoimmune disease is associated with altered *FDFT1* gene expression in immune cells, we performed a differential analysis of *FDFT1* expression in immune cells from SLE patients, MS patients, RA patients, and healthy subjects using single-cell or bulk RNA-seq data (GSE138266(84), E-GEAD-397(85)). Consistent with our chromatin-based MS V2G results and the eQTL co-localization analyses, *FDTF1* is significantly upregulated in CD4+ T cells from MS patients, as well as in naïve B cells, circulating GCB cells (plasmablasts), NK cells, monocytes, and Treg (**Figure 4E**). Similarly, *FDFT1* is significantly upregulated in naïve and memory B cells from SLE patients, as well as in innate immune cell types, memory CD4+ T cells, Th1, Th2, Th17, TFH and Treg (**Figure 4F**). The RA and SLE variants are located in the first intron of the *BLK* gene encoding the B cell Src family tyrosine kinase, therefore we also performed a differential analysis of *BLK* expression. *BLK* was expressed highly in B cell subsets and exhibited differential expression in SLE patients, but unlike *FDFT1*, *BLK* was expressed at low levels in T cells and was not differentially expressed in memory or differentiated CD4+ T cells from patients vs. controls (**Figure S15**).

To test whether *FDFT1* regulates inflammatory T cell function, we used the squalene synthase inhibitor lapaquistat, a drug historically used to treat hypercholesterolemia, to target *FDFT1* in an *in vitro* human tonsillar organoid model of T cell-dependent germinal center B cell differentiation and humoral immunity(86) (**Figure 5A**). After 7 days in culture, T-B interactions in untreated control organoids supported the differentiation naïve and memory T helper cells into PD1+CXCR5+ TFH and the differentiation of naïve and memory B cells into germinal center B cells and CD27+CD38+ plasmablasts capable of producing high-affinity class-switched antibodies (**Figure 5B**). While the FDFT1 drug lapaquistat had no effect on the viability of any B cell or T cell subset (**Figure 5C**), it showed dose-dependent inhibition of GCB differentiation and particularly abrogated plasmablast differentiation (**Figure 5D** and **E**). As a consequence, lapaquistat treatment resulted in a major decrease in the secretion of IgM and class-switched IgG and IgA antibodies by these follicular organoids (**Figure 5F**).

**Figure 5.**
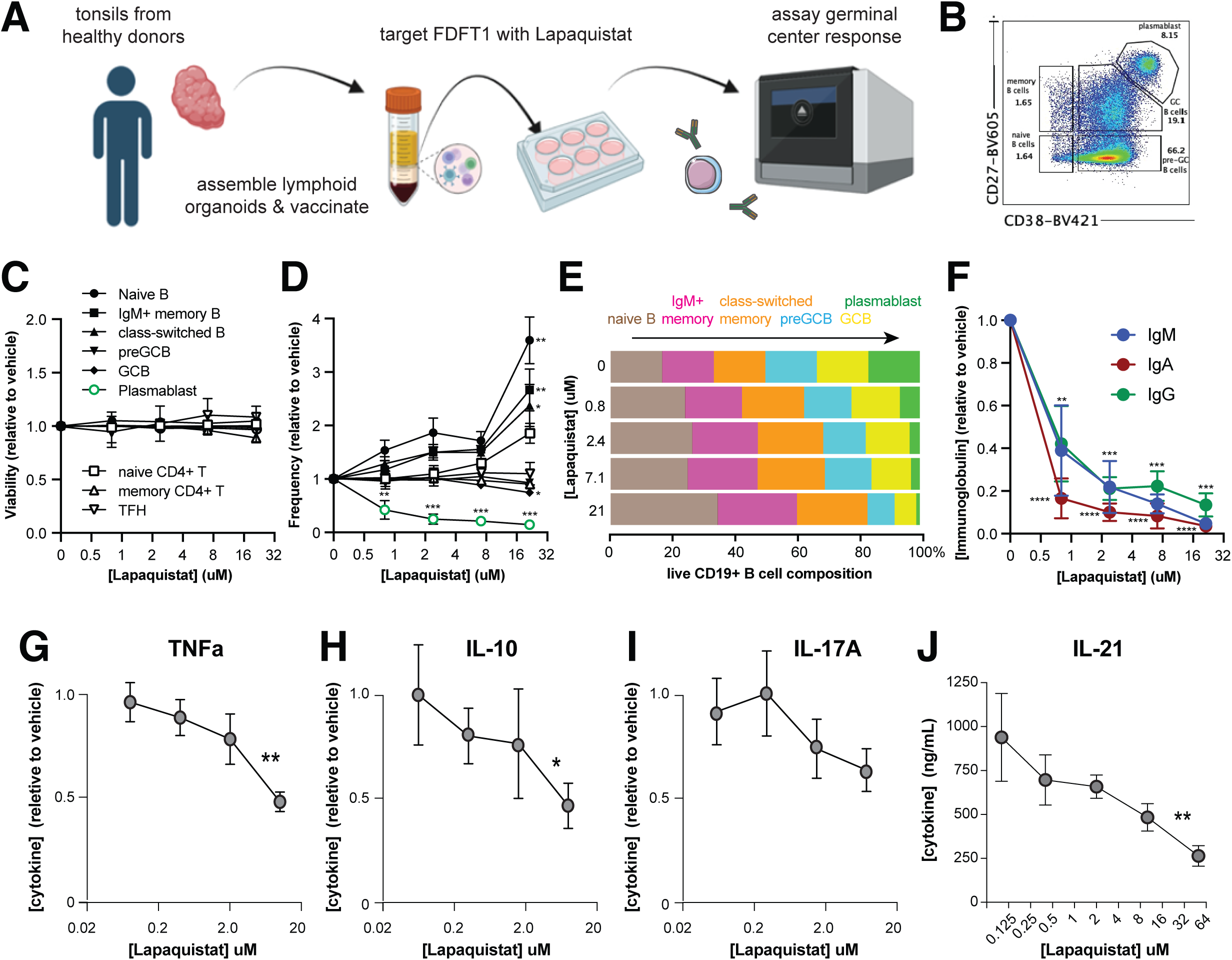
Targeting FDFT1 in an *in vitro* model of human T cell-dependent humoral immunity. Tonsillar organoid preparation, vaccination, functional read out scheme (**A**) and B cell gating strategy (**B**). Effect of the FDFT1 inhibitor lapaquistat on the viability (**C**) and frequency (**D**) of naïve B cells, memory B cells, class-switched B cells, germinal center B cells (GCB), plasmablasts, naïve CD4+ T cells, memory CD4+ T cells, and follicular helper T cells (TFH). B cell subset frequencies as in **D** but depicted as a proportion of all B cells across the B cell differentiation pathway (**E**). Effect of lapaquistat on immunoglobulin M, G, and A secretion (**F**) by the same tonsillar organoids as in **C-E**. N=5 donor tonsils; P values in **D** and **F** are: ****, <0.001, ***, <0.005, **, <0.01 and * <0.05. Effect of lapaquistat on activation-induced secretion of TNF (**G**), IL-10 (**H**), IL-17A (**I**), and IL-21 (**J**) by human tonsillar CD4+ T cells. N=5 tonsil donors; P values in **G-J** are: **, <0.01 and * <0.05.

Lapaquistat treatment induced a specific block in the GCB to plasmablast differentiation step that completely depends upon T cell help, indicating that squalene synthase activity may promote B cell antibody production by regulating T cell helper function. To test this, we isolated CD4+ T cells from human tonsil as a rich source of differentiated TFH and Th17 cells, stimulated them in the presence vs. absence of lapaquistat, and measured the secretion of TFH/Th17 cytokines known to support B cell antibody production and have important roles in the immunopathology characteristic of MS and SLE. Inhibition of squalene synthase activity had no effect on T cell activation at lower doses (**Figure S16**), but resulted in a dose-dependent decrease in activation-induced production of TNF and IL-17A, as well as decreased secretion of IL-10 (**Figure 5G-I**). The effect of lapaquistat on TNF and IL-10 levels was statistically significant. We also measured secretion of IL-21, a cytokine produced by human TFH and Th17 cells that is crucial for antibody class switching and production by B cells. Lapaquistat treatment resulted in a statistically significant, dose-dependent reduction in IL-21 secretion by tonsillar helper T cells (**Figure 5J**), consistent with its negative effect on plasmablast differentiation and immunoglobulin secretion. These data validate our V2G mapping approach and establish a previously unappreciated role for *FDFT1* in the regulation of CD4+ T cell-mediated inflammation and humoral immunity

## DISCUSSION

Deciphering the complex interplay between genetic risk factors, cell types, and immune dysregulation is crucial for understanding the pathogenesis of autoimmune disease, and such insights have the potential to revolutionize diagnostics and treatments tailored for improved patient outcomes. Our multi-omic approach provided 3D maps of gene cis-regulatory architecture that powered LDSC analyses to identify the cell types most likely impacted by autoimmune disease-associated genetic variation, followed by identification of those active variants in specific cell types and mapping them to the genes they likely regulate. These results implicated different immune cell types in distinct autoimmune diseases, as well as some non-immune cell types like enteroids, neuronal cells, and pancreatic cells in diseases such as IBD and MS.

Multiple cell types exhibited negative heritability enrichment for several autoimmune diseases. However, these events were not persistent across those cell types or traits. Thus, it is unlikely the negative enrichments have resulted from misclassification of the alleles in the trait summary statistics, or from particularly biased genetic regions of interest in the cell types. Given that this phenomenon was more pronounced in AS, JIA and PSO, it is likely that the effect of relatively few variants due to sample size employed in the given GWAS contributed to statistical power limitations with the estimation of LDSC, resulting in negative regression slopes in LDSC models.

The convergence of functional disease-specific variants on the same genes and biological networks across different cell types revealed by our 3D chromatin-based variant-to-gene mapping suggests that autoimmune diseases, despite their diverse manifestations, might share common pathways and mechanisms of influence. This places emphasis on the need for better predictive diagnostics with changes in specific gene expression potentially resulting from multiple autoimmune conditions. Understanding that different autoimmune diseases may converge on the same genes and networks implies that therapies targeting these shared pathways could be effective across multiple diseases. This opens up the possibility of developing broad-spectrum treatments that can address the root causes of various autoimmune disorders simultaneously. Conversely, the aspect of cell type-trait specific variants provides the opportunity for precision, personalized medicine, where therapies can be designed to modulate and target only a specific cell type of interest.

We also compared our 3D chromatin-based effector gene identification approach to eQTL approaches. We find that only 20-30% of the nominated genes were implicated by both 3D chromatin V2G and GTEx. The GTEx immune-related dataset is limited to whole blood, and is therefore not powered to determine which individual cell types are driving any of the detected eQTL. Approximately 30% of the nucleated cells in human whole blood are lymphocytes (B cells, T cells, NK cells), ∼5% are monocytes, and ∼60% are neutrophils, therefore neutrophils, a cell type not included in our analyses, are likely a major driver of GTEx immune eQTLs. The DICE and OneK1K scRNA-seq, and eQTL-Catalogue and cell type-specific RNA-seq datasets identified eQTL in immune cell types comparable to those included in our study, however, only 4.5% of the eGenes nominated were implicated by both approaches. The limited overlap could be due in part to the fact that these datasets implicated less than one quarter the number of eGenes as our study, but crucially, is also consistent with a recent study by Mostafavi et al. suggesting that eQTL and GWAS variants have evolved to control different types of loci(87).

The genes implicated by contact with autoimmune-associated variants include several that are part of known disease biology, as well as many that have not been studied in the context of the immunity or autoimmunity. These genes could represent novel therapeutic targets for controlling inflammatory disease. One such gene that we chose to follow up on is *FDFT1*, encoding a squalene synthase in the cholesterol pathway, which was captured in contact with an MS-associated variant in TFH cells and Th17 cells. While this enzyme has been studied extensively in the context of neuronal myelination, little is known about its specific role in immune cells. We show that FDFT1 antagonism using a direct, small molecule inhibitor of squalene synthase activity reduced production of IL-17, IFNg, and TNF by activated human CD4+ T cells. We did not establish the mechanism by which FDFT1 contributes to pro-inflammatory cytokine production in this study, however, a potential link to IL-17 is that the nuclear receptor RORgt, which is required for Th17 cell differentiation and function, is activated by oxysterols derived from cholesterol catabolism, and presumably would depend on squalene synthase activity(88,89). The link to TNF and IFNg is not clear, but a recent in vivo CRISPR screen implicated *FDFT1* in the development of memory CD8+ T cells, which are major producers of TNF and IFNg. Other study found that *FDFT1* activity disfavors mitochondrial oxidative phosphorylation by diverting lipid synthesis away from CoQ toward cholesterol synthesis(90). While more research is needed, our data suggest that *FDFT1* is a novel immunomodulator, and validates the power of 3D chromatin-based autoimmune variant-to-gene mapping strategies for identifying therapeutic targets for immune disease.

This study’s comprehensive exploration of genetic correlations, tissue-specificity, and shared pathways provides invaluable insights into the complex landscape of autoimmune diseases. By elucidating the underlying mechanisms and highlighting avenues for further research and therapeutic development, the study contributes to advancing our understanding and treatment of autoimmune disorders.

**Table S1. Data resources of 57 cell types**

**Table S2. Immune cells donors and mAbs details**

**Table S3. 16 autoimmune disorders**

**Table S4. Details of 4 cell type-specific eQTL databases**

**Table S5. Full LDSC analysis result**

**Table S6. Full variant-to-gene mapping result**

**Table S7. Full KEGG pathway enrichment analysis result**

**Table S8. Full GO-term enrichment analysis result**

**Table S9. Details of 258 loci used for eQTL analysis**

**Table S10. Shared cell types specific and trait specific eGenes between eQTLs datasets and V2G**

**Table S11. High PP.H4.abf colocalization results with coloc.abf**

**Table S12. High PP.H3.abf colocalization hits results with coloc.susie**

**Table S13. Colocalization analysis of FDFT1 on locus chr8:9921161-11972641**

**Figure S1. <u>Full S-LDSC parameters across diverse cell types’ cREs annotation</u>. A**. Bar plots (left) is the same in Figure 1A. The dot plots depict heritability enrichment for each cell type across 16 autoimmune traits as determined by LDSC analysis. Whiskers represent enrichment standard errors, with colors matched for HiC vs. capture-C. The colors of the dots correspond to p-values in -log10, with dots featuring a white asterisk indicating a significant p-value ≤ 0.05. The size of the dots corresponds to the proportion of SNP contribution to heritability. A dashed line at 1 indicates no enrichment. **B**. Bar plots (left) is the same in Figure 1A. The bar plot depicts conditional effect sizes for each cell type’s cREs annotation across 16 autoimmune traits as determined by LDSC analysis. The gradient color of the bar corresponds to the p-value in -log10, with a framed bar indicating a significant p-value ≤ 0.05. Bars with red asterisks passed FDR ≤ 0.05.

**Figure S2. <u>Bar plots show number of significant (P-values < 0.05) heritability</u> <u>enrichment</u>**for cell types per trait **(A)** and for traits per cell type **(B)**

**Figure S3. <u>Bar plots show number of significant (P-values < 0.05) conditional effect sizes</u>** (whole bars) and FDR<0.05 (green portions) for cell types per trait **(A)** and for traits per cell type **(B).** Volcano plots show conditional effect sizes versus P-values for cell types per trait **(C)** and for traits per cell type **(D)**, red dashed line is threshold p-value=0.05

**Figure S4.** (A) <u>Upset-plot showing intersections of V2G genes implicated in any cell type across 16 immune traits. (B) Upset-plot depicting intersections of V2G genes for any trait across the cell types listed.</u>

**Figure S5. <u>Degree of overlap</u>** of implicated variants (A) and target genes (B) across cell types

**Figure S6. <u>Cytokine/receptor gene enrichment</u>** across trait and cell type (A) and in KEGG pathway (B)

**Figure S7. <u>Salmonella infection gene enrichment</u>** across trait and cell type (A) and in KEGG pathway (B).

**Figure S8. <u>Sharing of V2G genes in enteroids</u>** across UC, CRO, and IBD (A). Gene ontology (B) and pathway enrichment (C) across each trait are shown.

**Figure S9. <u>Gene ontology enrichment</u>** of cell type-specific V2G genes across cell type.

**Figure S10.** Shared eGenes across different eQTL datasets with V2G

**Figure S11.** Shared eGenes across different eQTL datasets per locus

**Figure S12. <u>Collage of pie plots each depicting the proportion of eGenes identified by</u> <u>eQTL</u>** that match (light orange) or do not match (purple) the gene identified by 3D chromatin V2G for each cell type-trait pairing. **Left panels**: bar plots depicting the total number of eQTL eGenes for each immune cell type that match or differ across all autoimmune traits. **Bottom panels**: bar plots depicting the total number of eQTL eGenes for each trait that match or differ across all immune cell types

**Figure S13. <u>Disrupted transcription factor binding motifs</u>**: Motifs of transcription factors where binding sites were predicted to be affected by high-probability causal variants (SNPs with PP.H4.abf ≥ 0.8) that overlapped with variants identified by the V2G (variant-to-gene) approach. Altered alleles are presented, consistent with the corresponding GWAS of the immune traits.

**Figure S14. <u>Dot-plot shows effect sizes of SLE and RA variants</u>** on *FDFT1* expression in different cell types, shaped according to the origin of datasets, colored by whether the variant were SLE leads or their proxies.

**Figure S15. <u>Expression of BLK</u>** in sorted immune cells from RA patients, SLE patients, and healthy subjects measured by bulk RNA-seq. Statistically significant differential expression compared to healthy subjects is denoted by p-values.

**Figure S16. <u>Effect of lapaquistat on T cell activation</u>** as measured by induction of IL-2 receptor (A) and IL-2 (B) expression and proliferation (C) by human CD4 T cells stimulated with anti-CD3+CD28 beads.

## Data Availability

All data and supplementary tables from this study are available upon reasonable request to the authors and will be published following peer-reviewed acceptance.

## ACKNOWLEDGMENTS

S.F.A.G. is the Daniel B. Burke Endowed Chair for Diabetes Research.

