## Supplementary figures S1-S16 for "3D chromatin-based variant-to-gene maps across 57 human cell types reveal the cellular and genetic architecture of autoimmune disease susceptibility"

**Figure S1** - Full S-LDSC parameters across diverse cell types' cREs annotation. A. Bar plots (left) is thesame in Figure 1A. The dot plots depict heritability enrichment for each cell type across 16 autoimmune traits as determined by LDSC analysis. Whiskers represent enrichment standard errors, with colors matched for HiC vs. capture-C. The colors of the dots correspond to p-values in -log10, with dots featuring a white asterisk indicating a significant p-value ≤ 0.05. The size of the dots corresponds to the proportion of SNP contribution to heritability. A dashed line at 1 indicates no enrichment. B. Bar plots (left) is the same in Figure 1A. The bar plot depicts conditional effect sizes for each cell type's cREs annotation across 16 autoimmune traits as determined by LDSC analysis. The gradient color of the bar corresponds to the p-value in -log10, with a framed bar indicating a significant p-value ≤ 0.05. Bars with red asterisks passed FDR ≤ 0.05.

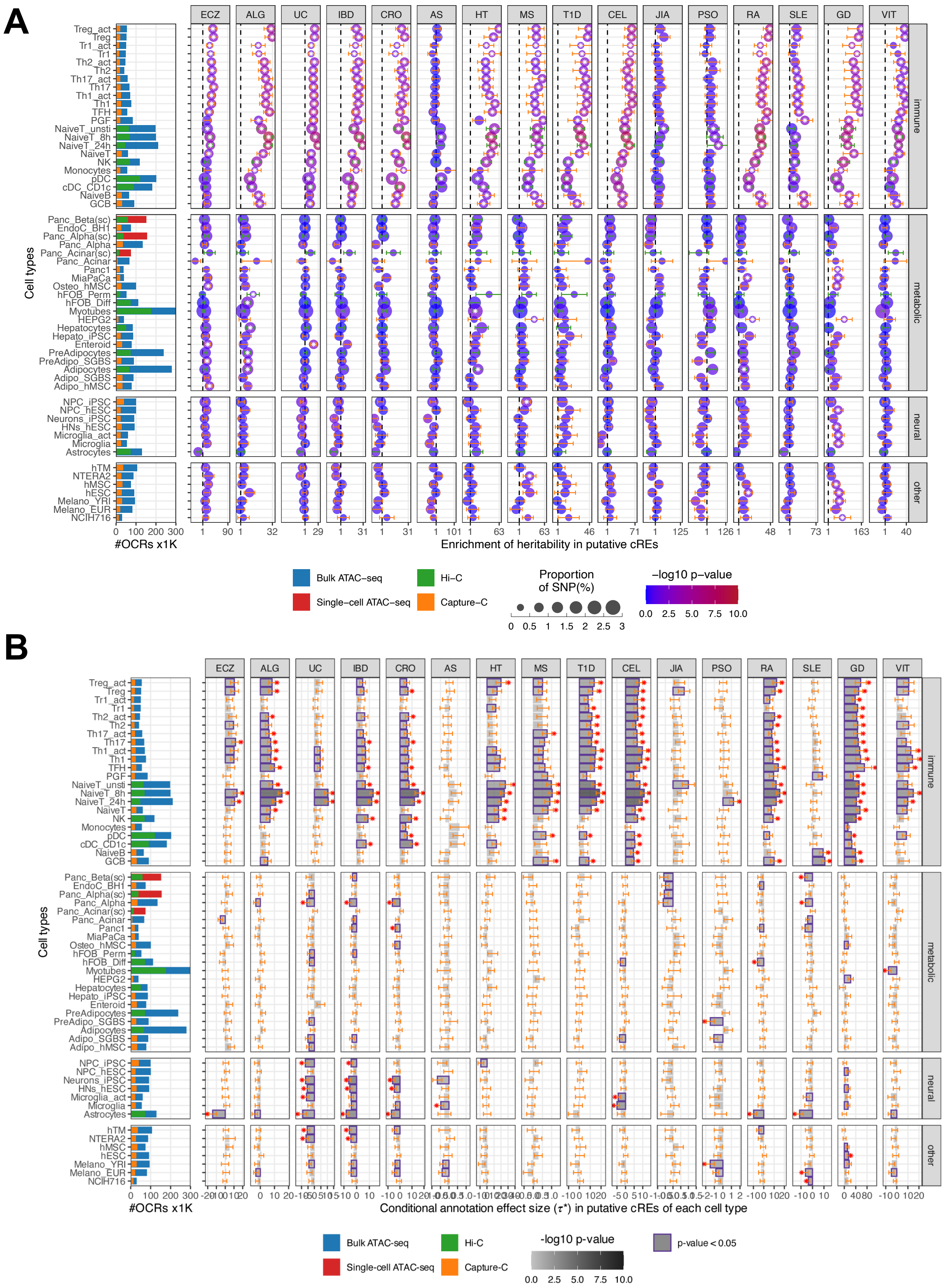

**Figure S2 -** Plots show number of significant (P<0.05) heritability enrichment for cell types per trait (A) and for traits per cell type (B).

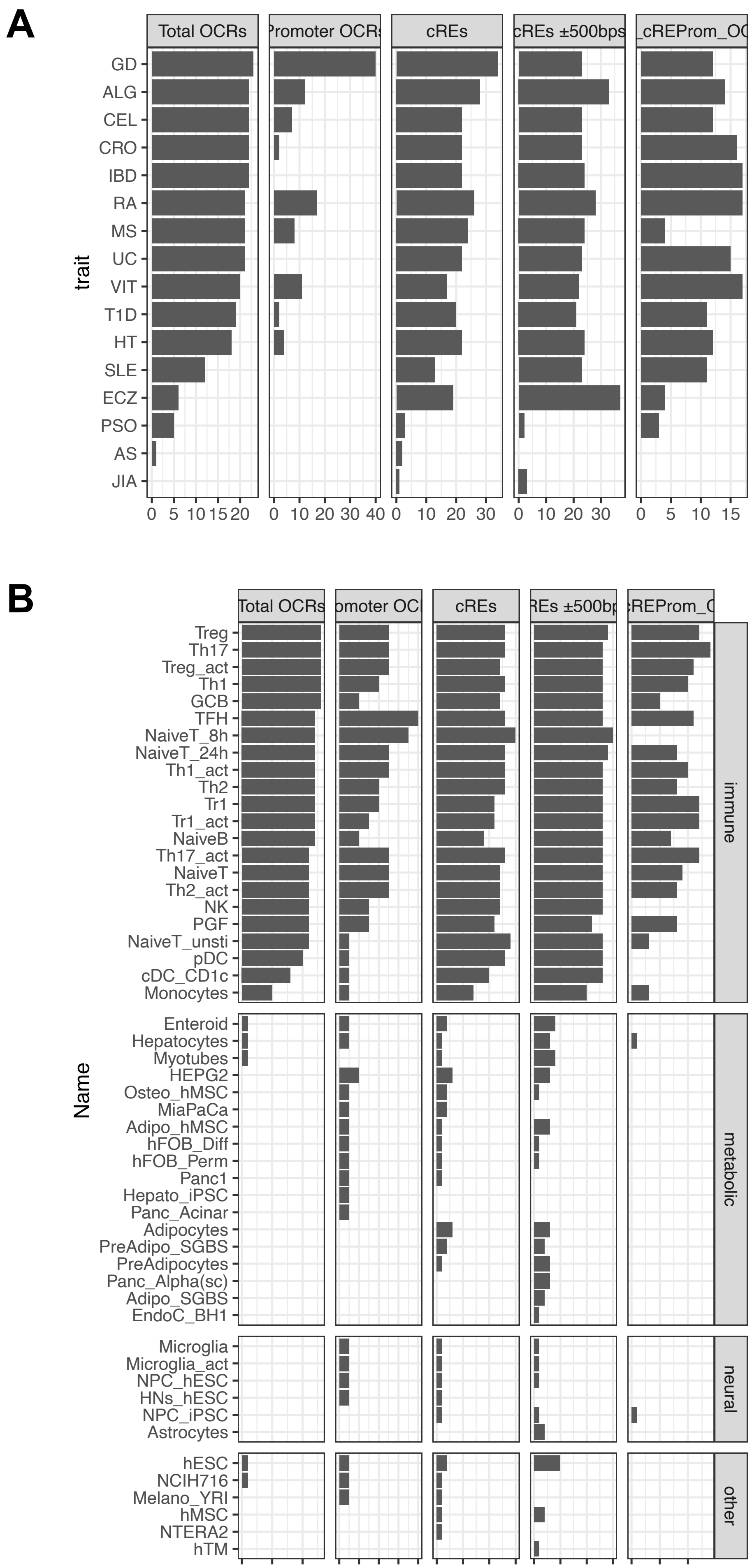

**Figure S3 -** Bar plots show number of significant (P-values < 0.05) conditional effect sizes (whole bars) and FDR<0.05 (green portions) for cell types per trait (A) and for traits per cell type (B). Volcano plots show conditional effect sizes versus P-values for cell types per trait (C) and for traits per cell type (D), red dashed line is threshold p-value=0.05.

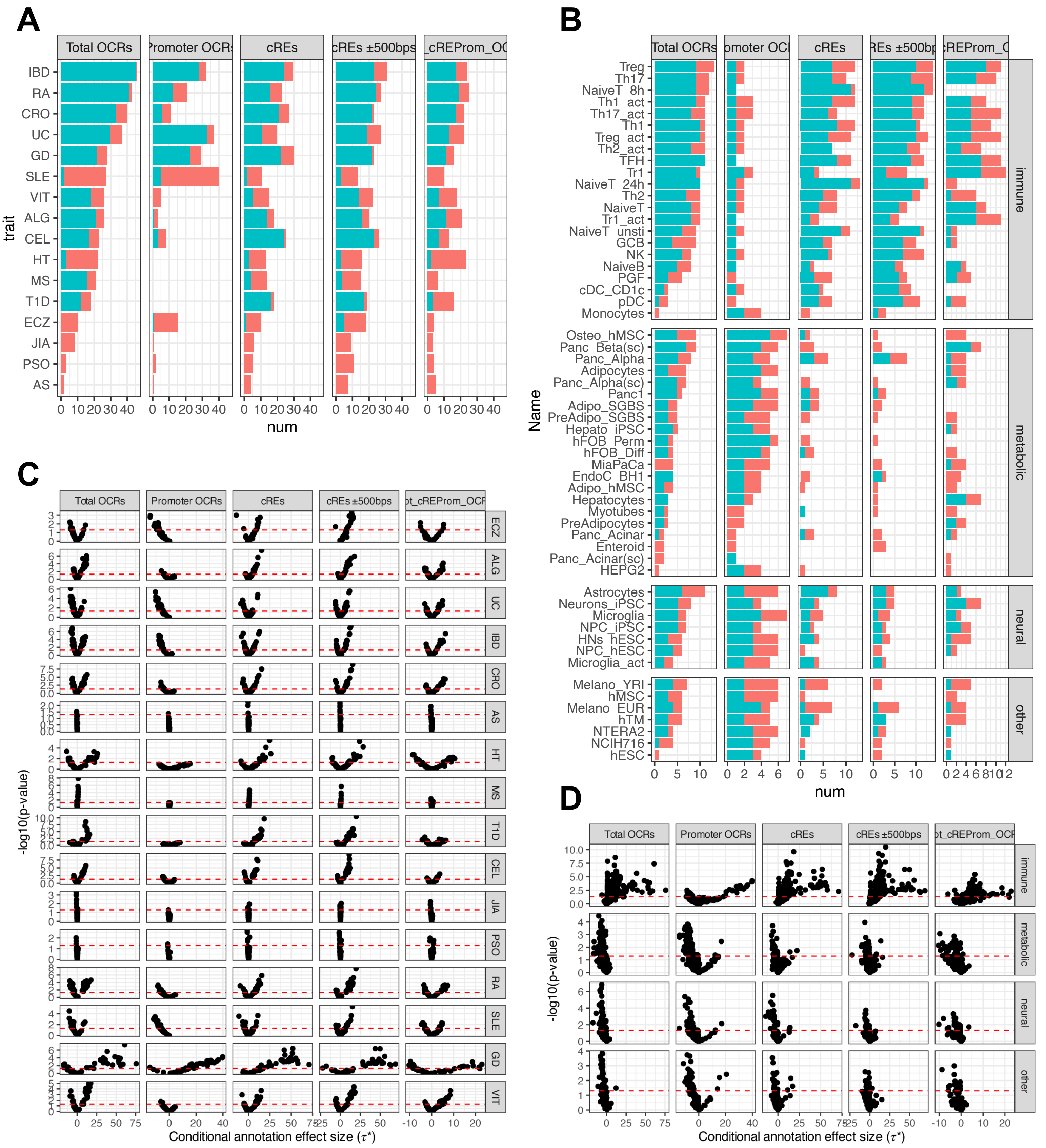

**Figure S4 -** (A) Upset-plot showing intersections of V2G genes implicated in any cell type across 16 immune traits. (B) Upset-plot depicting intersections of V2G genes for any trait across the cell types listed.

**A**

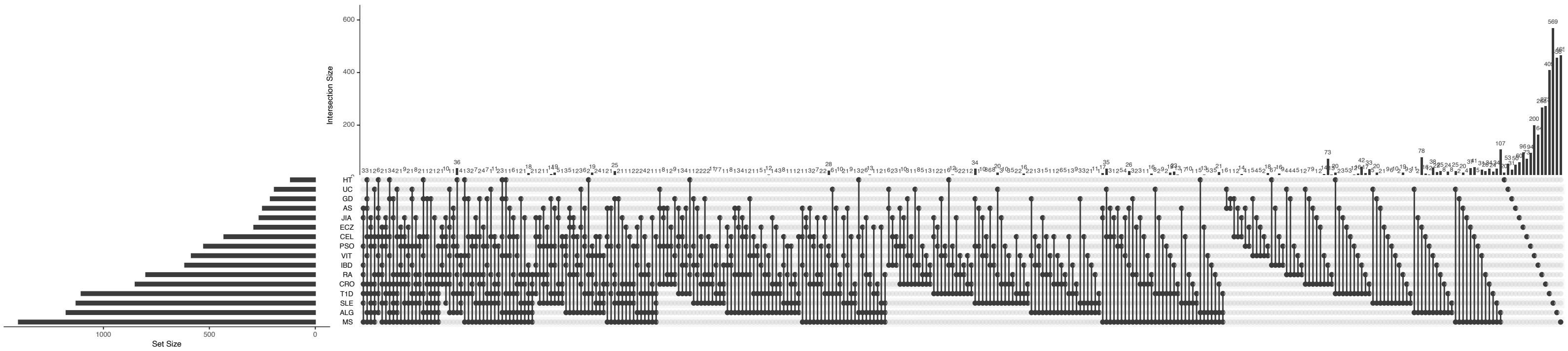


**B**

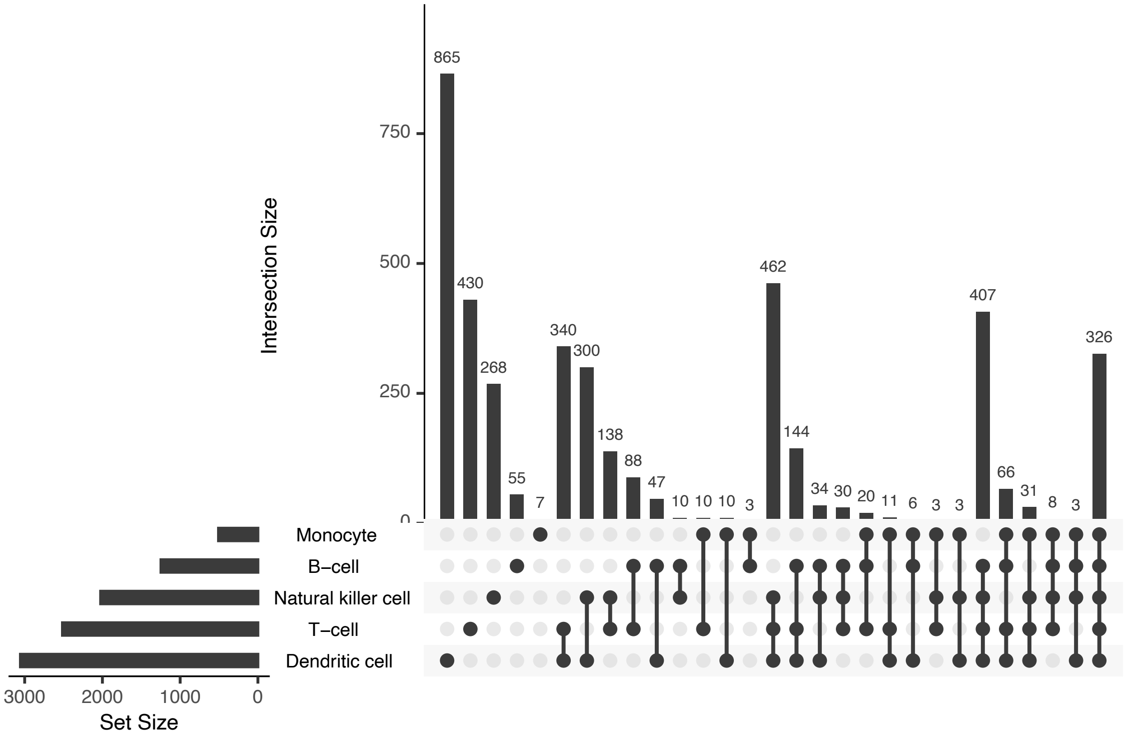

**Figure S5 -** Degree of overlap of implicated variants (A) and target genes (B) across cell types

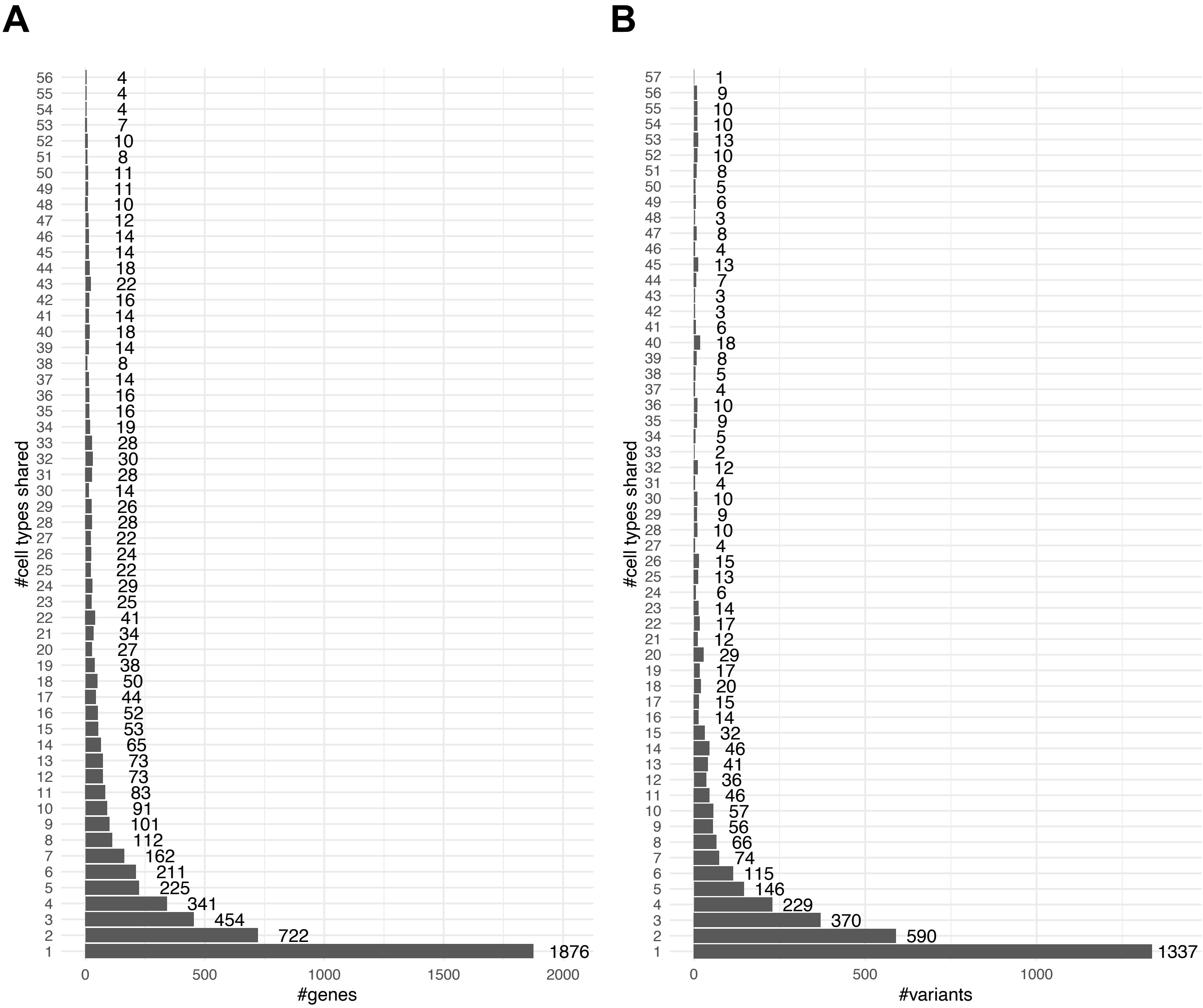

**Figure S6 -** Cytokine/receptor gene enrichment across trait and cell type (A) and in KEGG pathway (B)

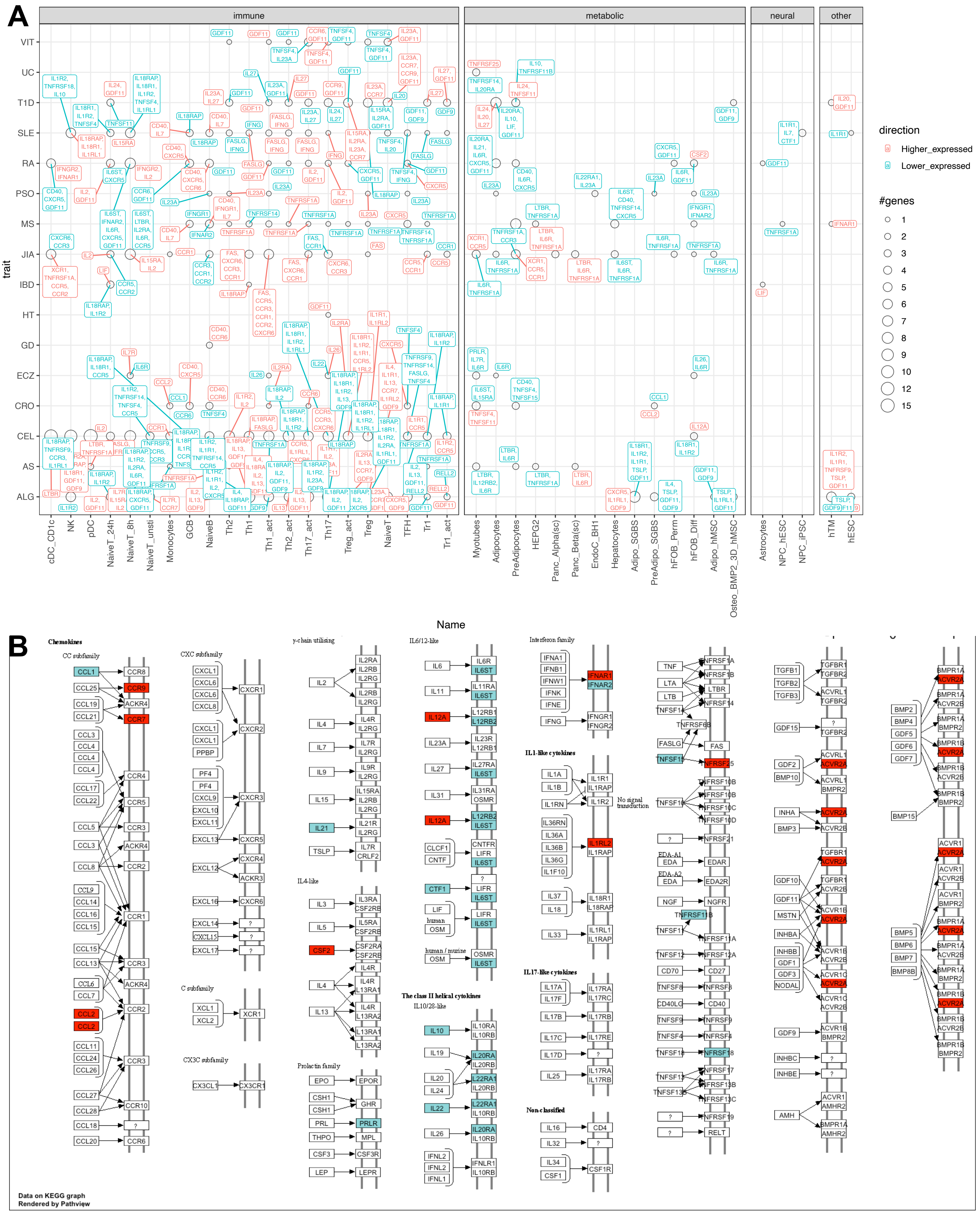

**Figure S7 -** Salmonella infection gene enrichment across trait and cell type (A) and in KEGG pathway (B).

A

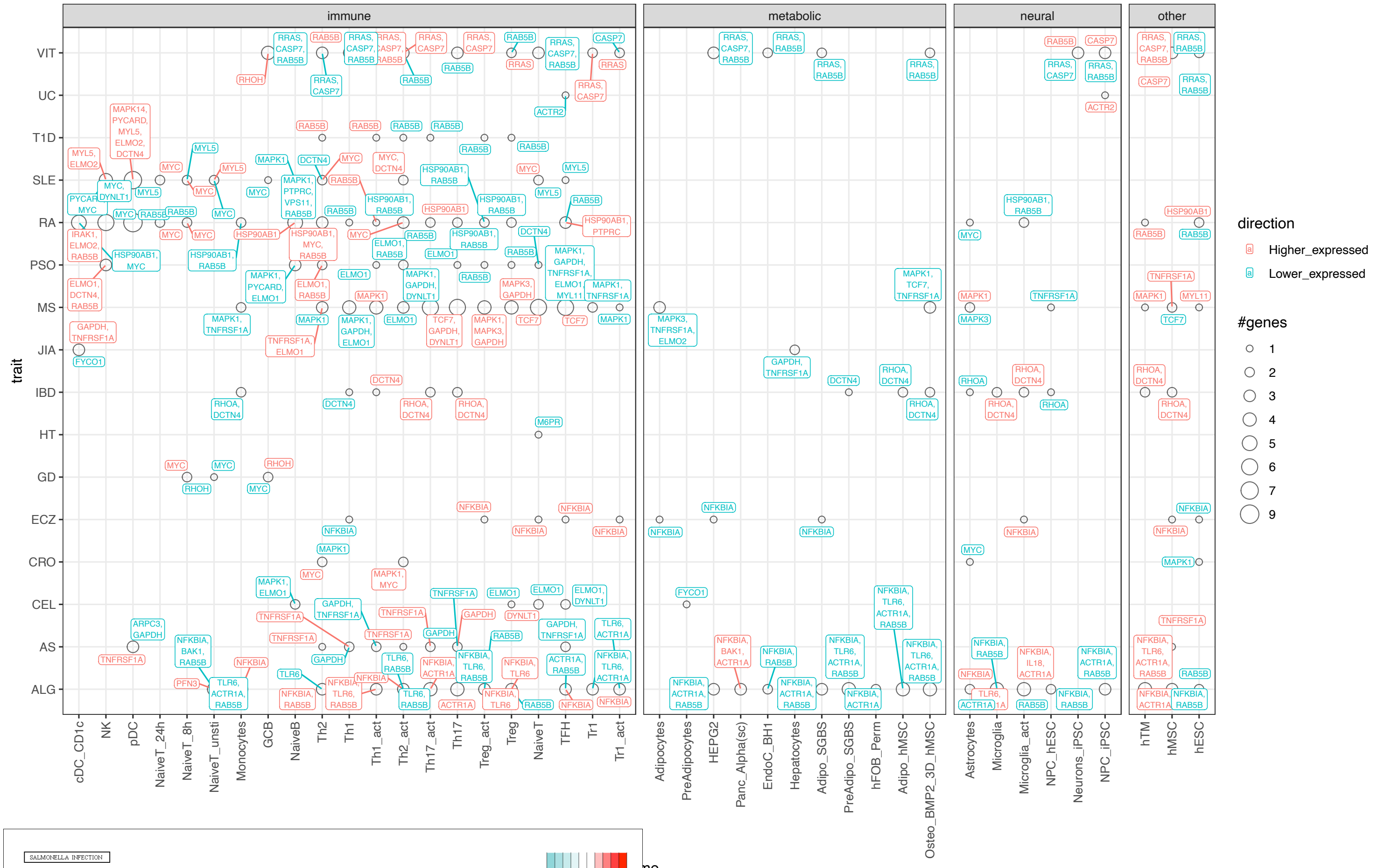


B

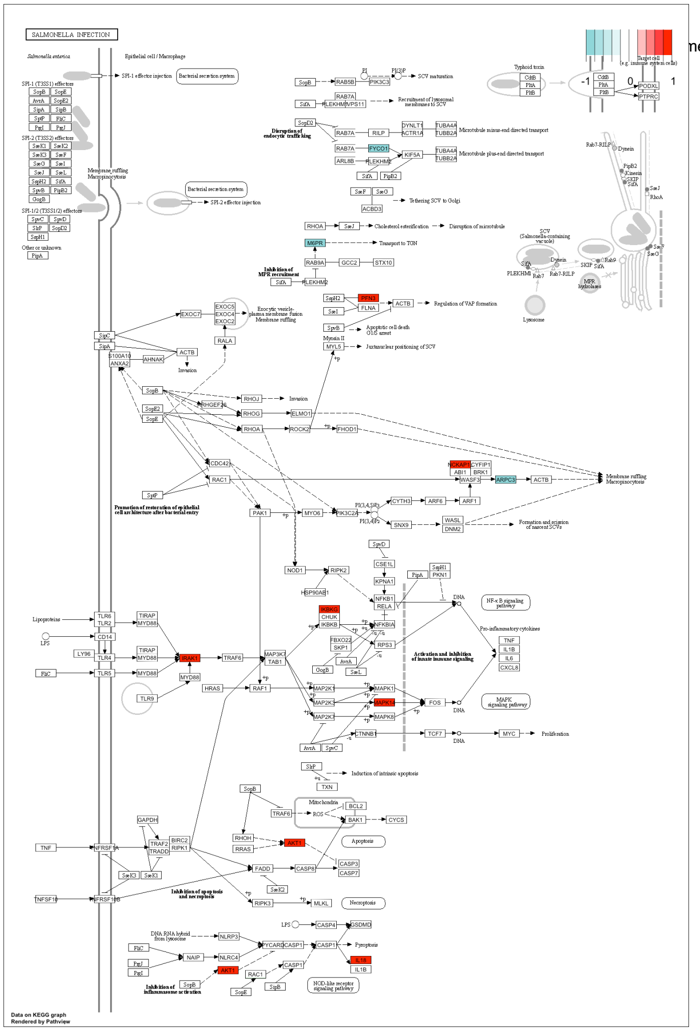

**Figure S8 -** Sharing of V2G genes in enteroids across UC, CRO, and IBD (A). Gene ontology (B) and pathway enrichment (C) across each trait are shown.

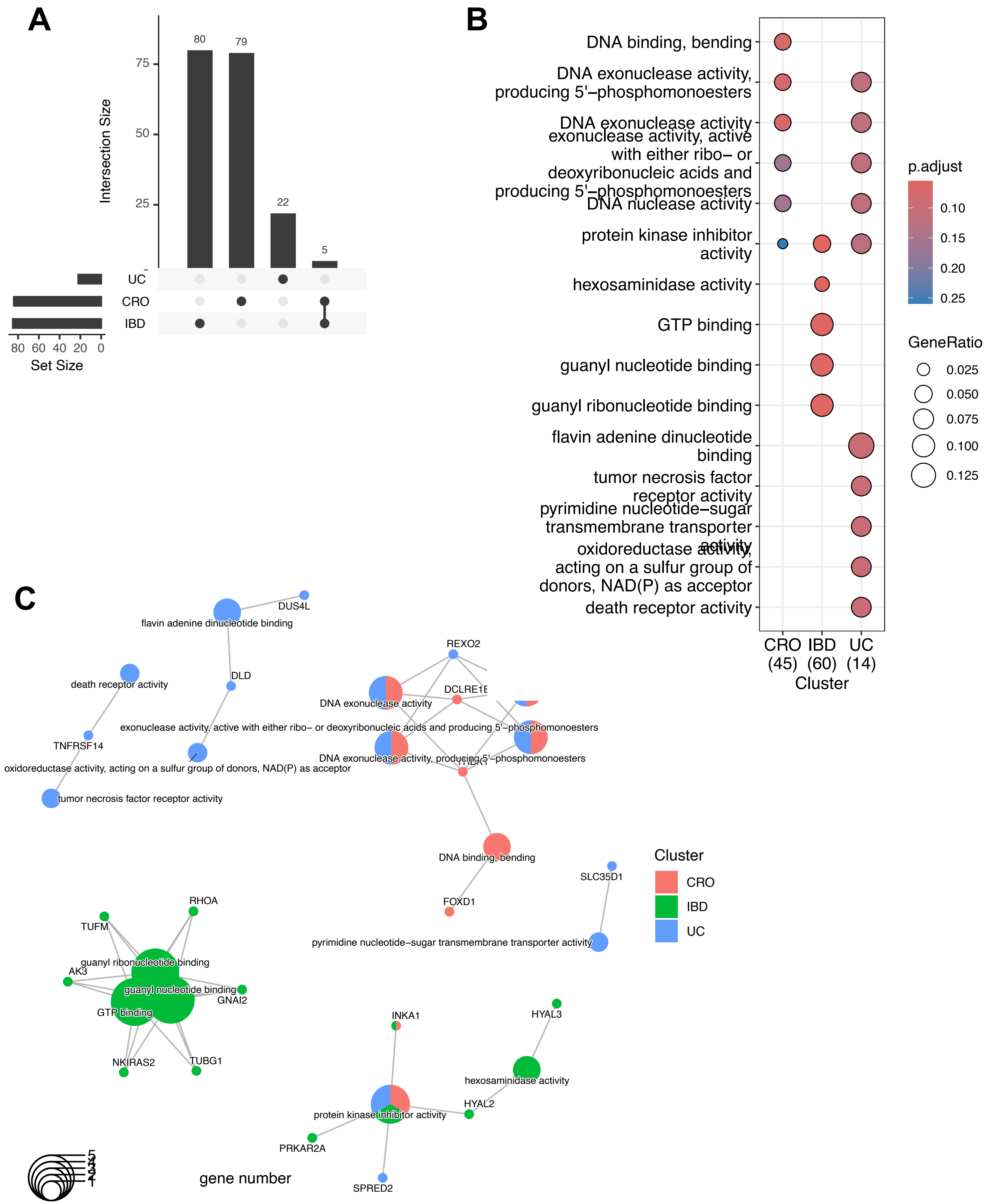

**Figure S9 -** Gene ontology enrichment of cell type-specific V2G genes across cell type.

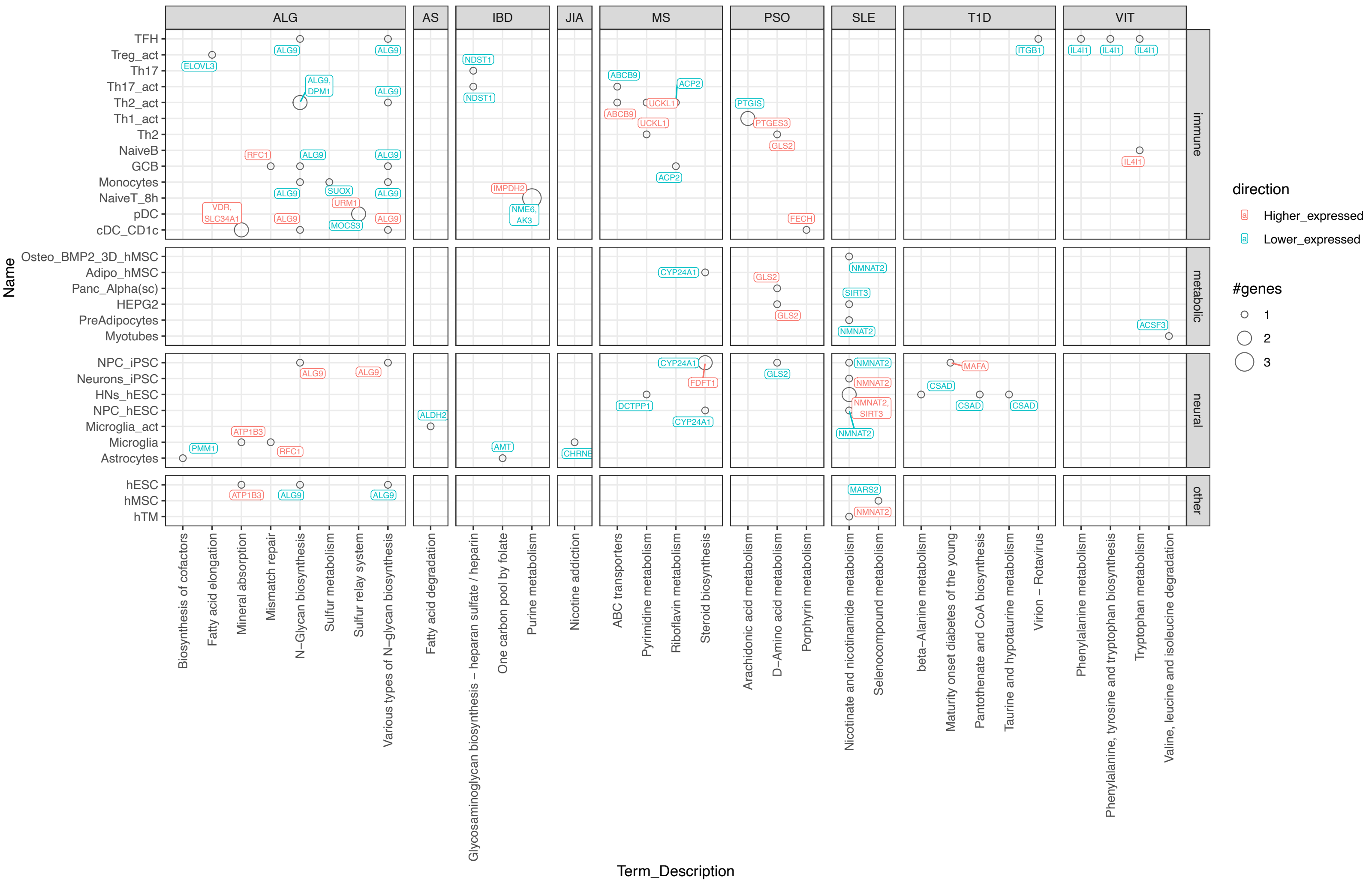

**Figure S10 -** Shared eGenes across different eQTL datasets.

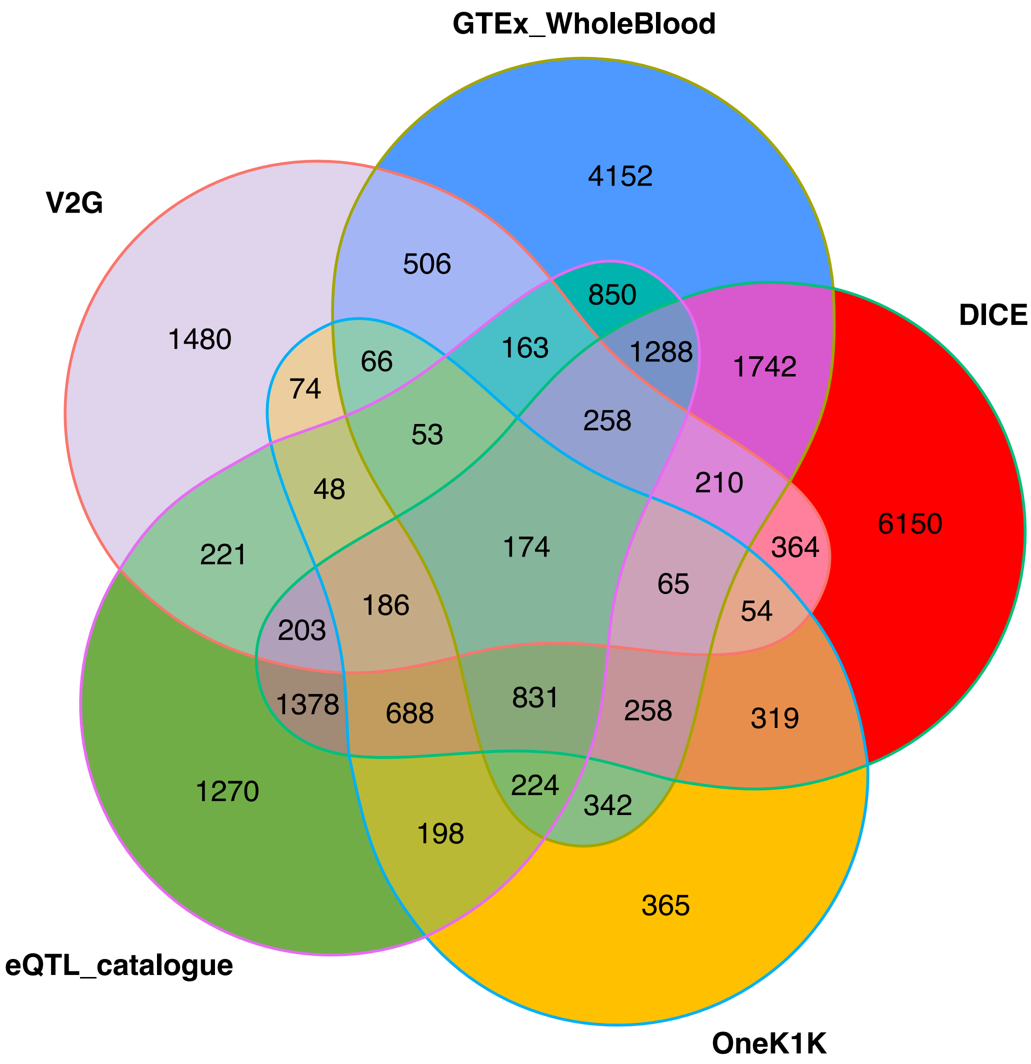

**Figure S11 -** Shared eGenes across different eQTL datasets with V2G

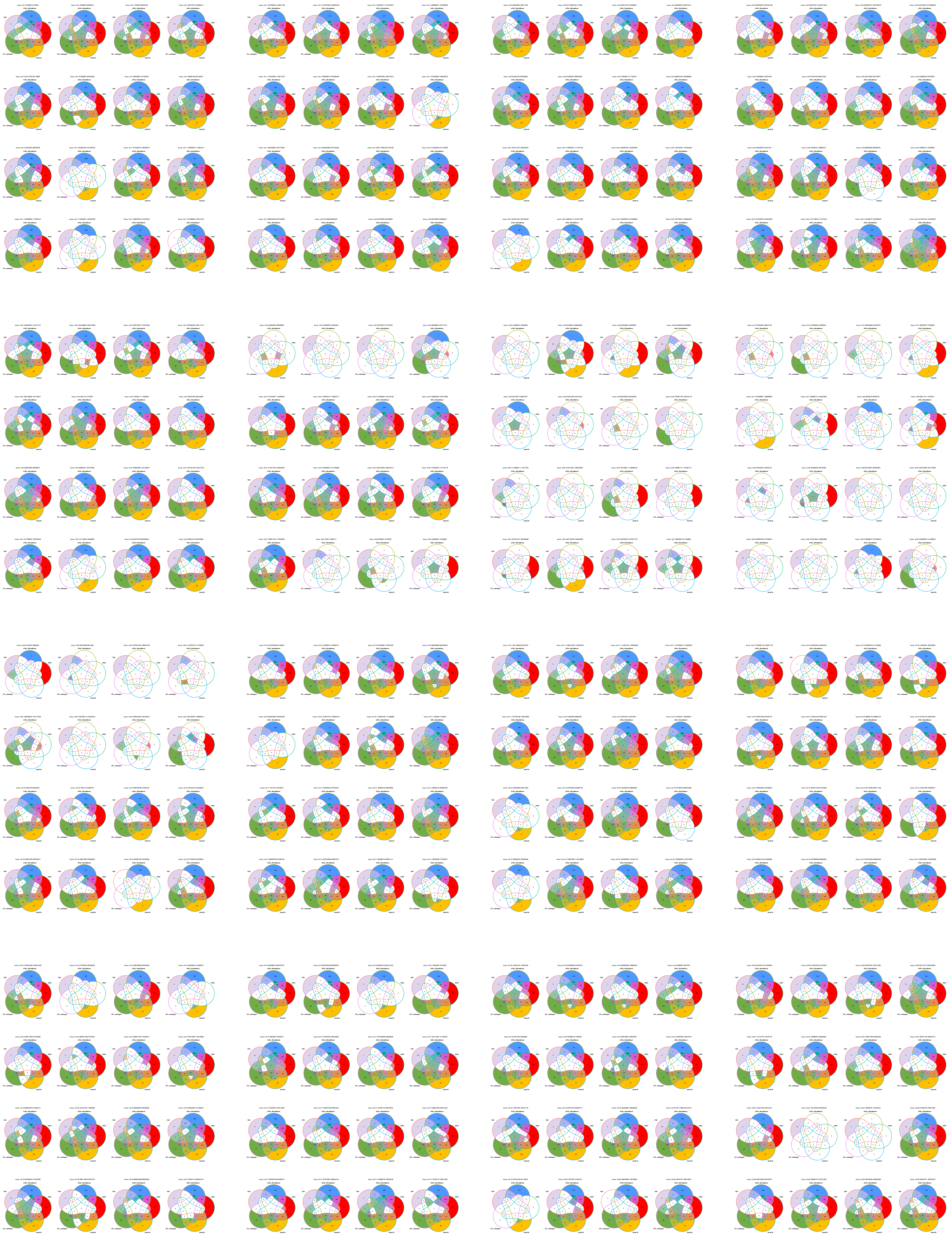

**Figure S12 -** Collage of pie plots each depicting the proportion of eGenes identified by eQTL that match (light orange) or do not match (purple) the gene identified by 3D chromatin V2G for each cell type-trait pairing. Left panels: bar plots depicting the total number of eQTL eGenes for each immune cell type that match or differ across all autoimmune traits. Bottom panels: bar plots depicting the total number of eQTL eGenes for each trait that match or differ across all immune cell types.

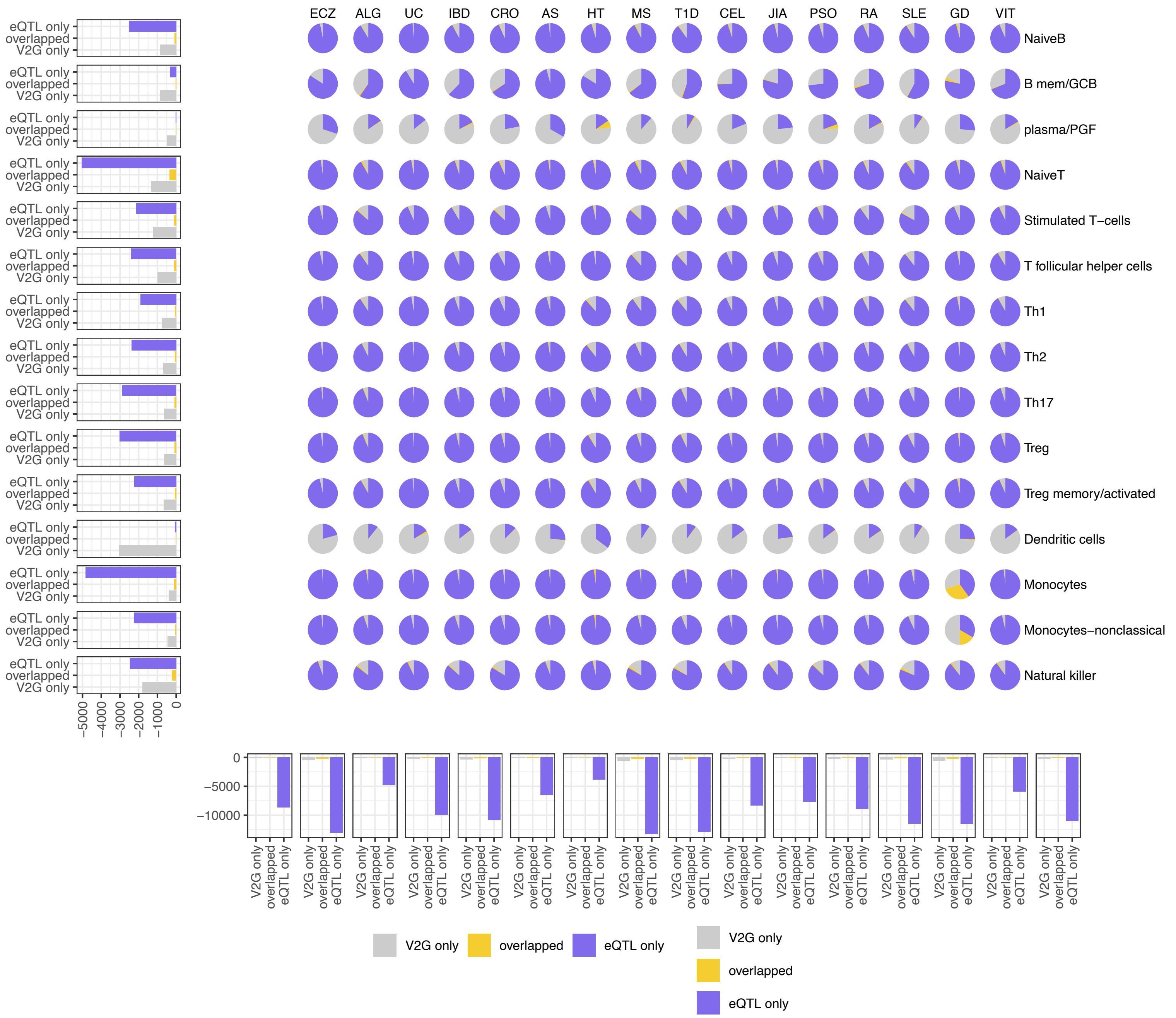

**Figure S13 -** Disrupted transcription factor binding motifs: Motifs of transcription factors where binding sites were predicted to be affected by high-probability causal variants (SNPs with PP.H4.abf ≥ 0.8) that overlapped with variants identified by the V2G (variant-to-gene) approach. Altered alleles are presented, consistent with the corresponding GWAS of the immune traits.

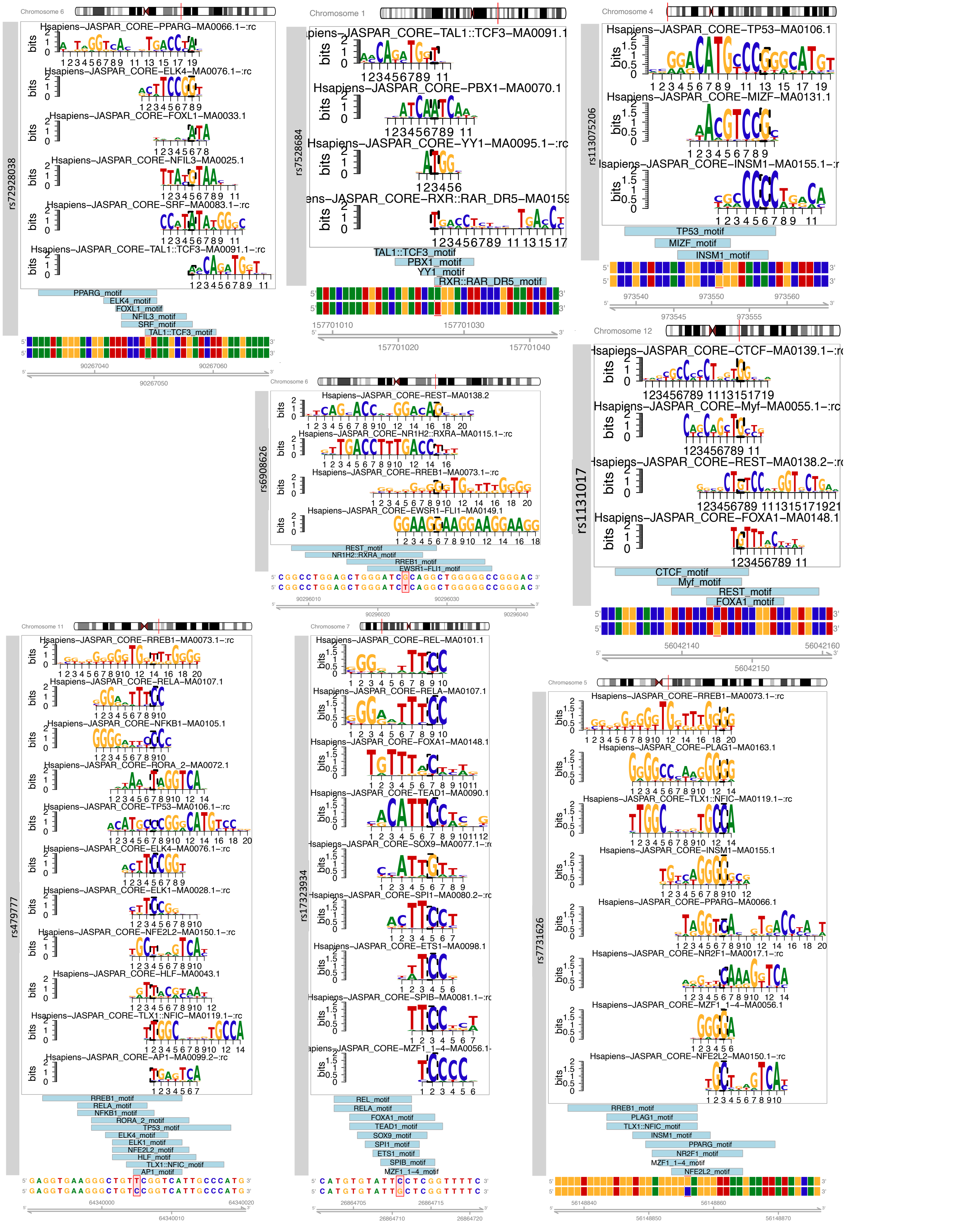

**Figure S14 -** Dot-plot shows effect sizes of SLE and RA variants on FDFT1 expression in different cell types, shaped according to the origin of datasets, colored by whether the variant were SLE leads or their proxies.

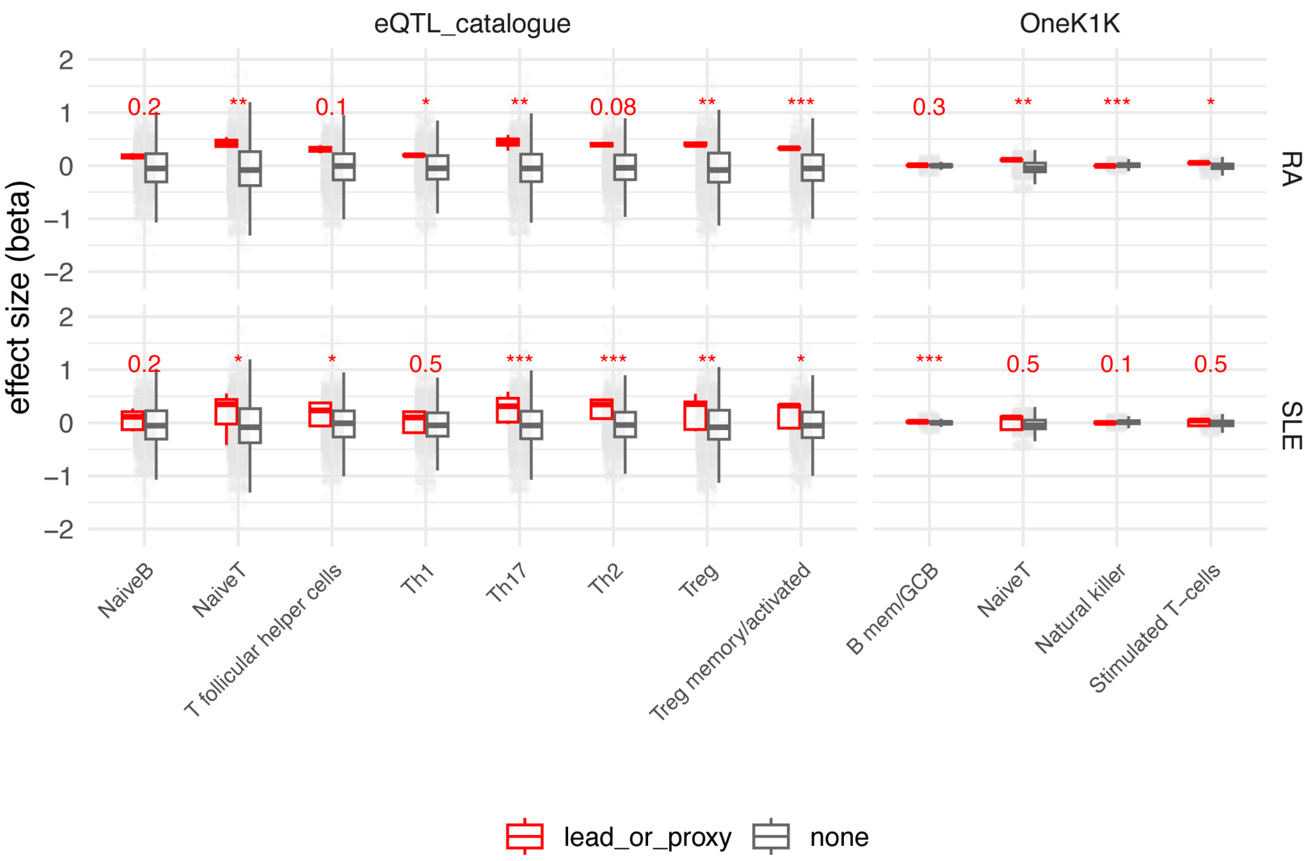

**Figure S15 -** Expression of BLK in sorted immune cells from RA patients, SLE patients, and healthy subjects measured by bulk RNA-seq. Statistically significant differential expression compared to healthy subjects is denoted by p-values.

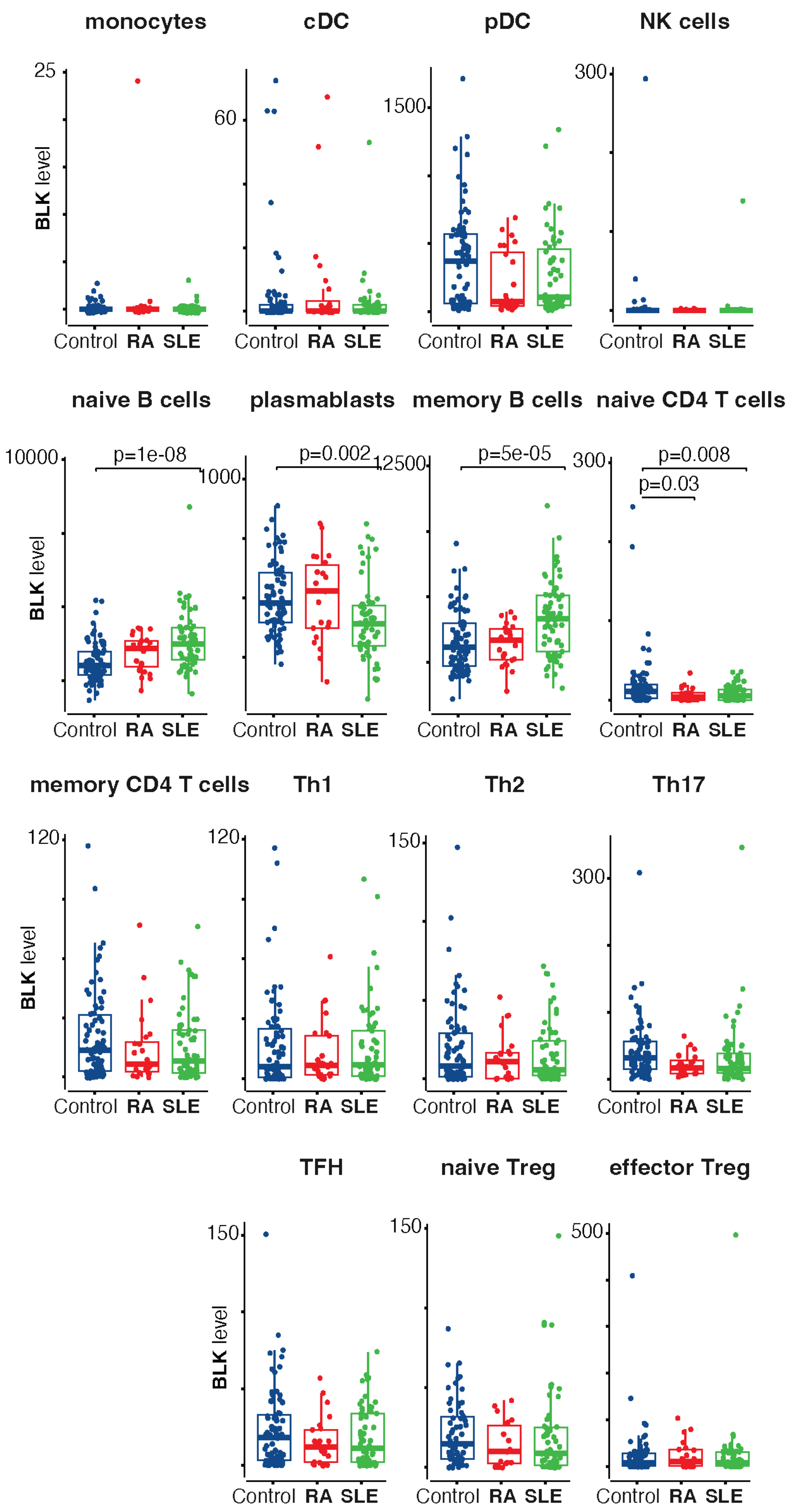

**Figure S16 -** Effect of lapaquistat on T cell activation as measured by induction of IL-2 receptor (A) and IL-2 (B) expression and proliferation (C) by human CD4 T cells stimulated with anti-CD3+CD28 beads.

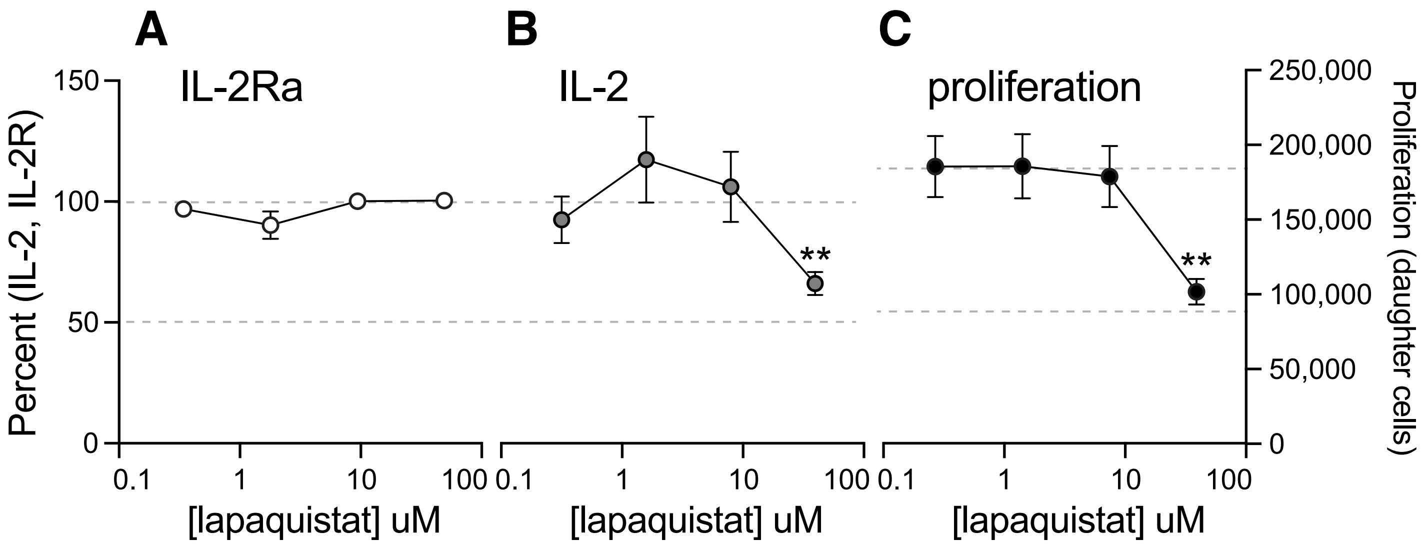
