## Supplemental File 1 for "3D chromatin-based variant-to-gene maps across 57 human cell types reveal the cellular and genetic architecture of autoimmune disease susceptibility"

### Supplementary File 1

#### Gating strategy for sorting of plasmacytoid dendritic cells (pDC) and CD1c+ dendritic cells (cDC2)

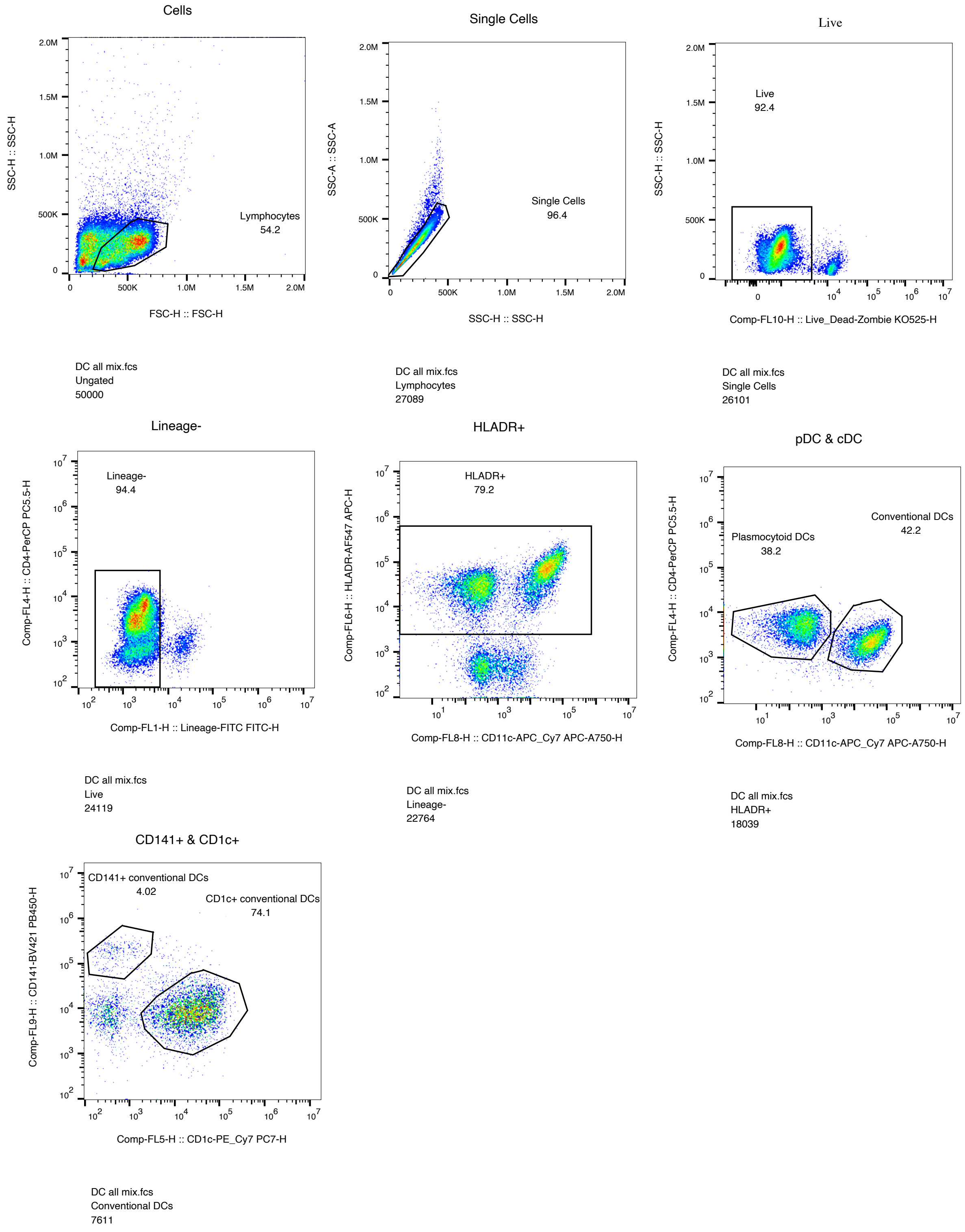

Gating strategy for sorting of Th1, Th2, Th17, Tr1, and Treg subsets

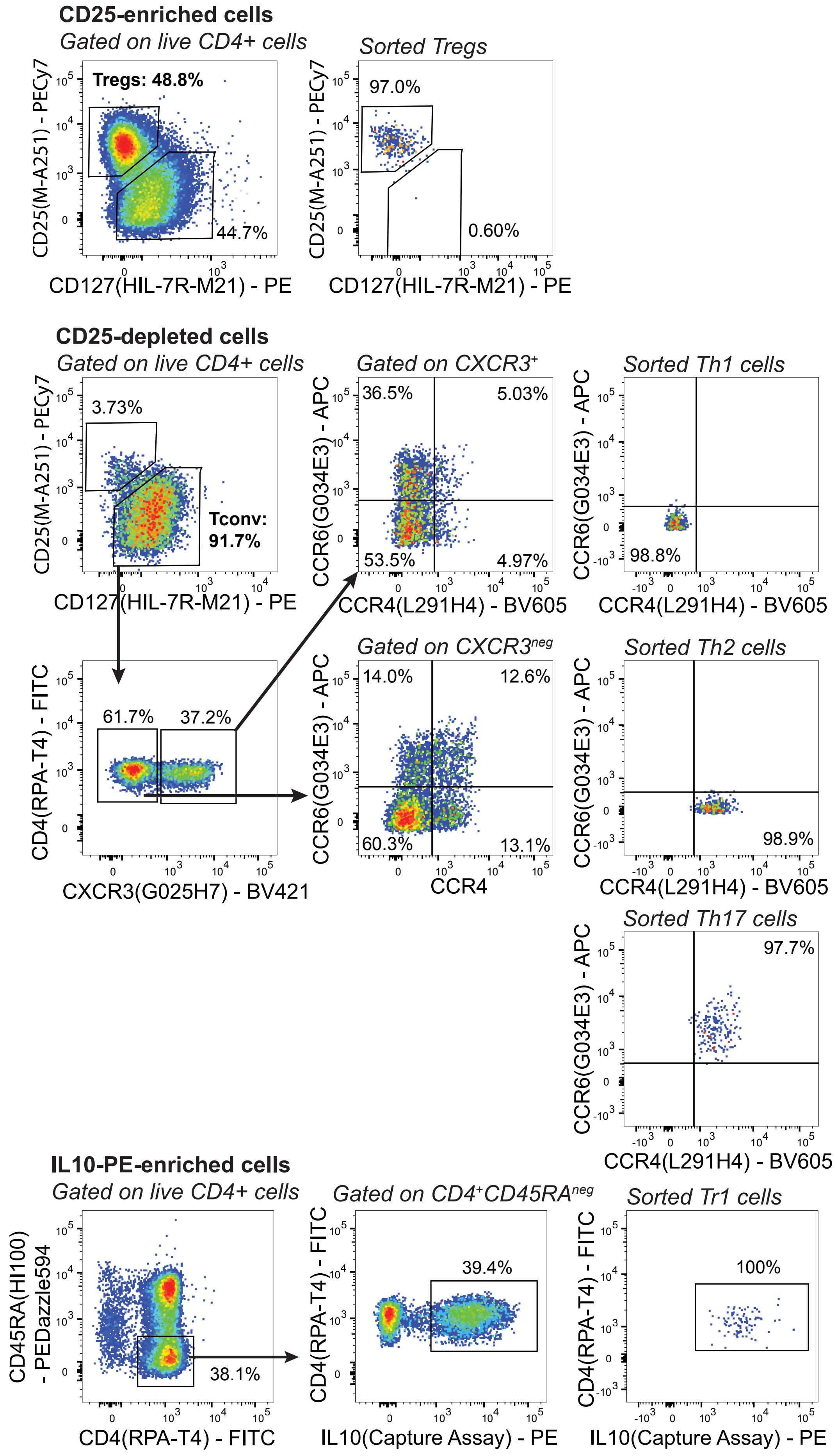

Phenotypic characterization of sorted Th1, Th2, Th17, Tr1, and Treg subsets. See Table S2 for donor info.

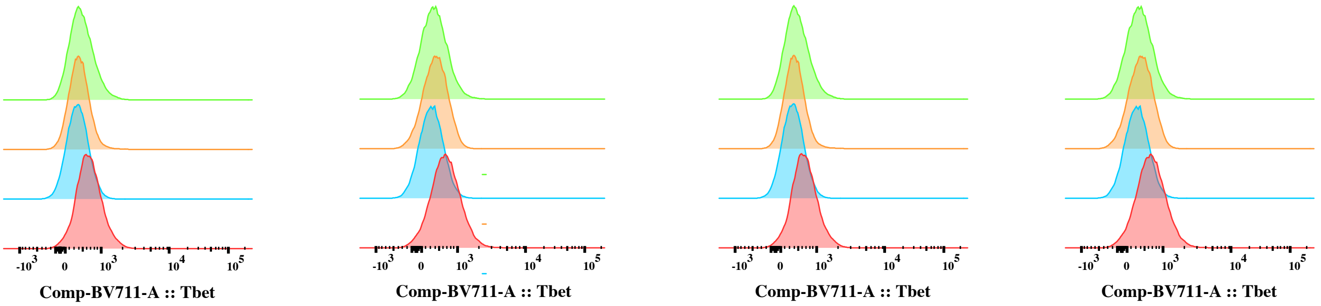


| | Sample Name | Geometric Mean : Comp-BV711-A |
|---|---|---|
| | 1700_Treg | 489 |
| | 1700_Th17 | 434 |
| | 1700_Th2 | 394 |
| | 1700_Th1 | 673 |

| | Sample Name | Geometric Mean : Comp-BV711-A |
|---|---|---|
| | 5549_Treg | 401 |
| | 5549_Th17 | 421 |
| | 5549_Th2 | 337 |
| | 5549_Th1 | 678 |

| | Sample Name | Geometric Mean : Comp-BV711-A |
|---|---|---|
| | 1700_Treg | 489 |
| | 1700_Th17 | 434 |
| | 1700_Th2 | 394 |
| | 1700_Th1 | 673 |

| | Sample Name | Geometric Mean : Comp-BV711-A |
|---|---|---|
| | 5549_Treg | 401 |
| | 5549_Th17 | 421 |
| | 5549_Th2 | 337 |
| | 5549_Th1 | 678 |

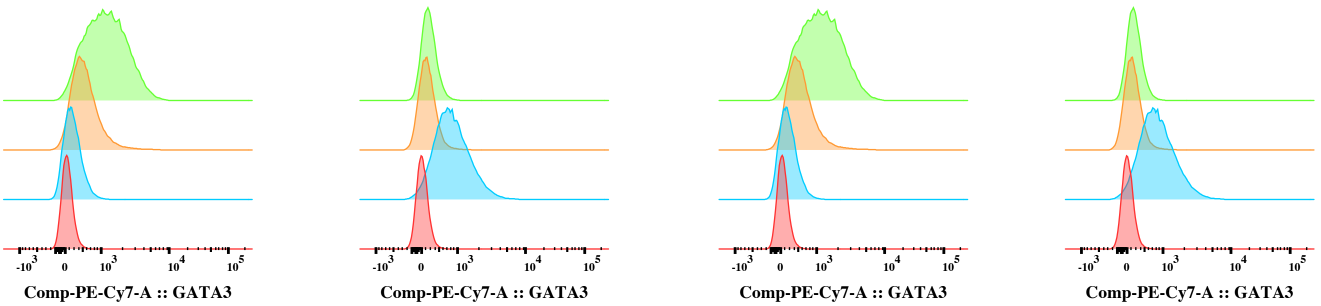


| | Sample Name | Geometric Mean : Comp-PE-Cy7-A |
|---|---|---|
| | 1700_Treg | 1201 |
| | 1700_Th17 | 572 |
| | 1700_Th2 | 284 |
| | 1700_Th1 | 168 |

| | Sample Name | Geometric Mean : Comp-PE-Cy7-A |
|---|---|---|
| | 5549_Treg | 284 |
| | 5549_Th17 | 249 |
| | 5549_Th2 | 864 |
| | 5549_Th1 | 141 |

| | Sample Name | Geometric Mean : Comp-PE-Cy7-A |
|---|---|---|
| | 1700_Treg | 1201 |
| | 1700_Th17 | 572 |
| | 1700_Th2 | 284 |
| | 1700_Th1 | 168 |

| | Sample Name | Geometric Mean : Comp-PE-Cy7-A |
|---|---|---|
| | 5549_Treg | 284 |
| | 5549_Th17 | 249 |
| | 5549_Th2 | 864 |
| | 5549_Th1 | 141 |

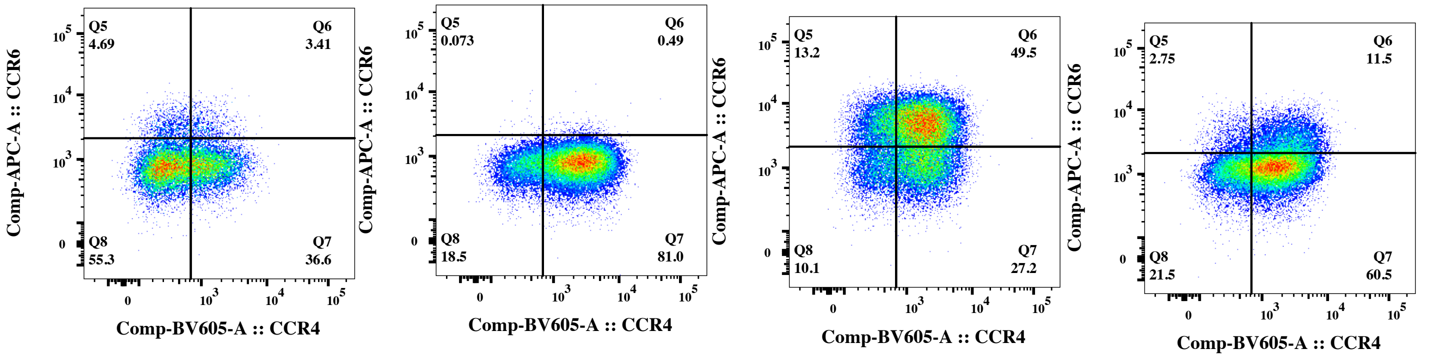


1700_Th1 1700_Th2 1700_Th17 1700_Treg

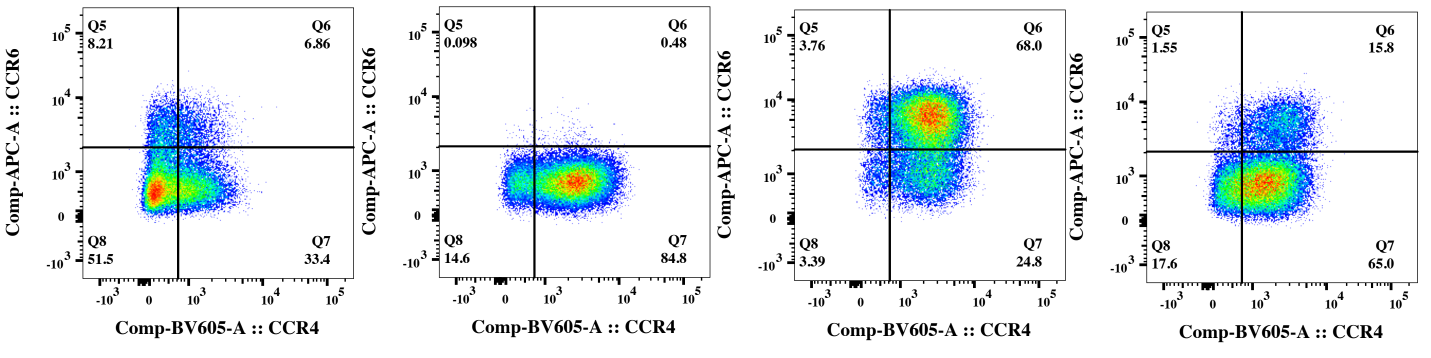


5549_Th1 5549_Th2 5549_Th17 5549_Treg

Phenotypic characterization of day 14 in vitro expanded Th1, Th2, Th17, Tr1 & Treg subsets.

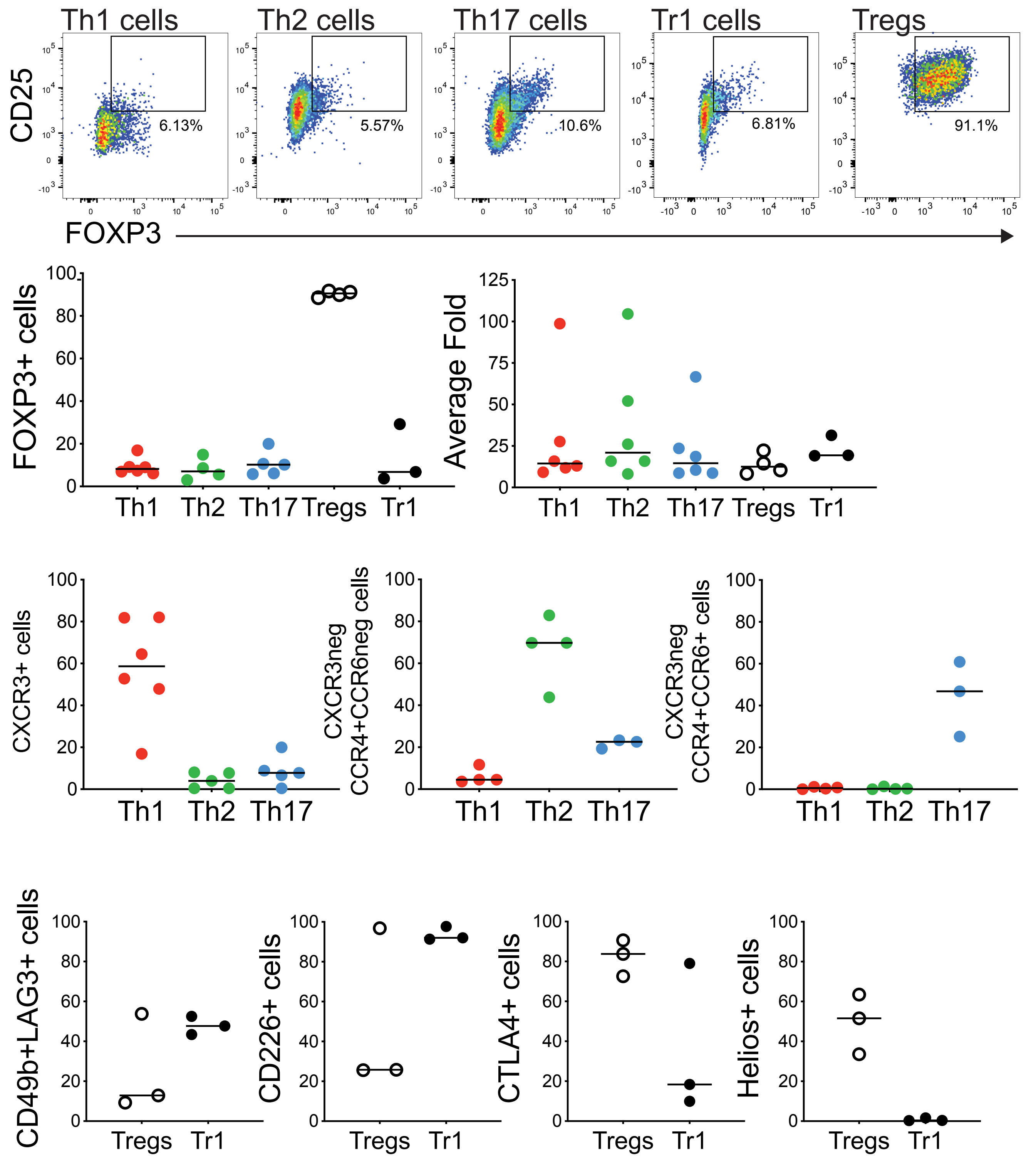

Functional characterization of day 14 in vitro expanded Th1, Th2, and Th17 subsets.

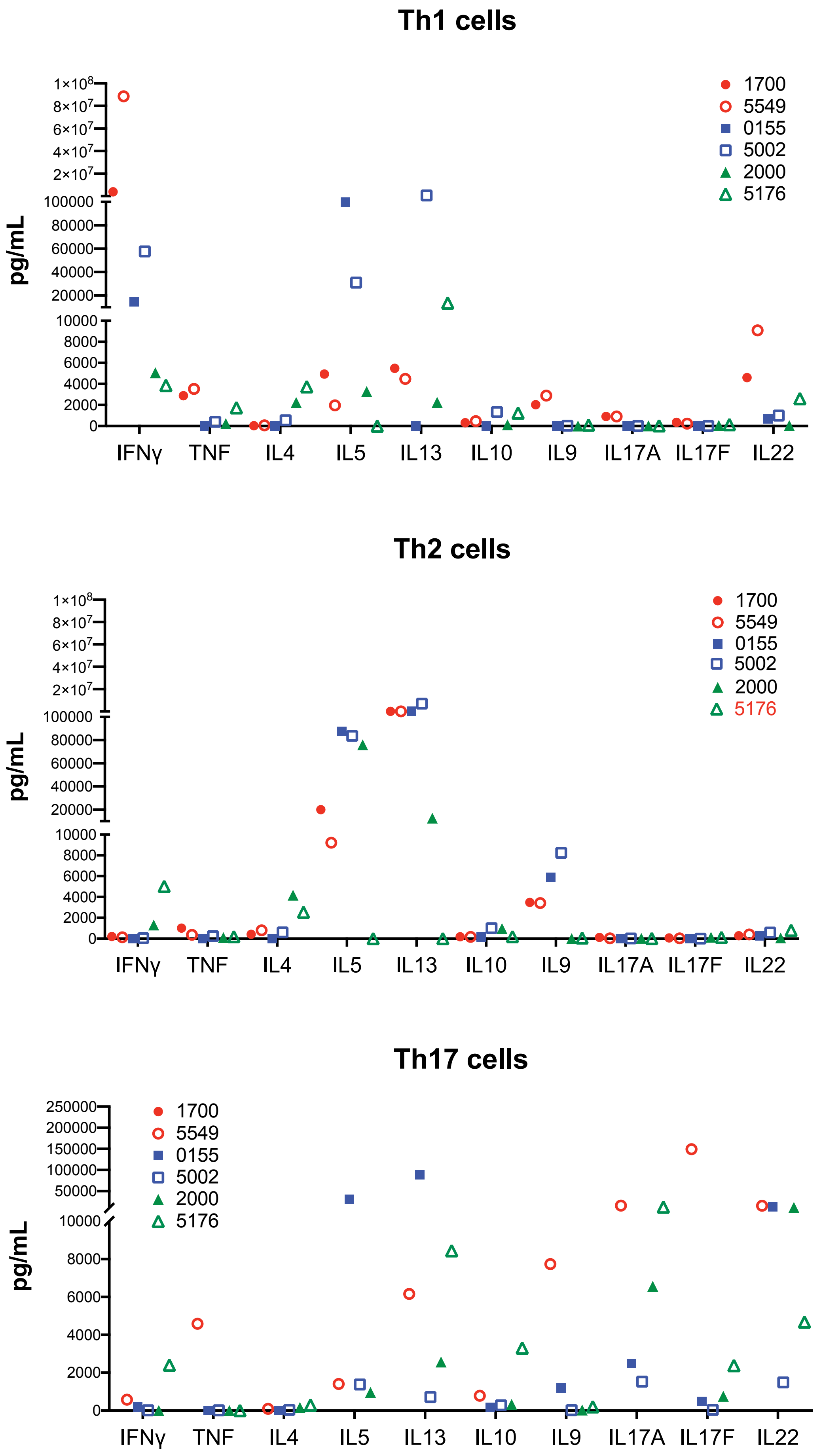
